# Impact of heat on respiratory hospitalizations among older adults living in 120 large US urban areas

**DOI:** 10.1101/2024.05.22.24307126

**Authors:** Cassandra R. O’Lenick, Stephanie E. Cleland, Lucas M. Neas, Mallory W. Turner, E. Melissa Mcinroe, K. Lloyd Hill, Andrew J. Ghio, Meghan E. Rebuli, Ilona Jaspers, Ana G. Rappold

**Affiliations:** University of North Carolina at Chapel Hill; School of Medicine; Chapel Hill, NC; United States Environmental Protection Agency; Clinical Research Branch; Chapel Hill, NC; University of North Carolina at Chapel Hill; Gillings School of Public Health; Department of Environmental Sciences and Engineering; Chapel Hill, NC; Simon Fraser University, Faculty of Health Sciences, Burnaby, British Columbia, Canada

**Keywords:** Extreme heat, Medicare, time-series, climate change epidemiology, respiratory hospitalizations

## Abstract

**Objectives:** A nationwide study of the impact of high temperature on respiratory disease hospitalizations among older adults (65+) living in large urban centers.

**Methods:** Daily rates of short-stay, inpatient respiratory hospitalizations were examined with respect to variations in ZIP-code-level daily mean temperature in the 120 largest US cities between 2000-2017. For each city, we estimated cumulative associations (lag-days 0-6) between warm-season temperatures (June-September) and cause-specific respiratory hospitalizations using time-stratified conditional quasi-Poisson regression with distributed lag non-linear models. We estimated nationwide associations using meta-regression and updated city-specific associations via best linear unbiased prediction. With stratified models, we explored effect modification by age, sex, and race (Black/white). Results were reported as percent change in hospitalizations at high temperatures (95th percentile) compared to median temperatures for each outcome, demographic-group, and metropolitan area. Excess hospitalization rates were estimated for days above median temperatures.

**Results:** At high temperatures, we observed increases in the percent of all-cause respiratory hospitalizations [1.2 (0.4, 2.0)], primarily driven by an increase in respiratory tract infections [1.8 (0.6, 3.0)], and chronic respiratory diseases/respiratory failure [1.2 (0.0, 2.4)]. East North Central, New England, Mid-Atlantic, and Pacific cities accounted for 98.5% of the excess burden. By demographic group, we observed disproportionate burdens of heat-related respiratory hospitalizations among the oldest beneficiaries (85+ years), and among Black beneficiaries living in South Atlantic cities.

**Conclusion:** This study found robust impacts of high temperature on respiratory failure and chronic inflammatory and fibrotic diseases among older adults. The geographic variation suggests that contextual factors account for disproportionate burdens.

## INTRODUCTION

Exposure to extreme heat and humidity has a direct impact on human health and well-being (1–3), and is projected to result in a 370% increase in heat-related mortality among those 65 years and over by mid-century (2). Robust associations between heat and excess mortality among older adults are consistently reported in the literature. However, associations between heat and health care utilization, such as emergency department visits and hospitalizations, are often weaker with inconsistent findings across study locations (4–9). Previous studies of heat and respiratory morbidity have primarily focused on all-cause respiratory events or major categories of respiratory disease (chronic obstructive pulmonary disease, asthma, and respiratory tract infections), with little attention given to less common respiratory diseases. The impact of extreme heat on cause-specific morbidity is an important knowledge gap that must be addressed to optimize patient outcomes and increase the resilience of health care systems for future extreme heat events.

Older adults are among the most physiologically sensitive to prolonged heat exposure because of the effects of aging on thermoregulation and evaporative heat loss. In addition, older adults may have impaired mobility, pre-existing diseases, or take medications that impact body fluid balance or interact with hemodynamic regulation (3, 10). When prolonged heat exposure results in hyperthermia, older adults may experience myocardial strain due to increased cardiac output, interstitial fluid pooling, as well as systemic inflammation and reactive oxygen species production (3, 11–13). Hyperthermia can also lead to increases in ventilation rate, tidal volume, and respiratory rate (14), which may damage the lung parenchyma and exacerbate underlying respiratory illnesses. Epidemiologic and mechanistic studies provide evidence that some heat-related pulmonary injury may also be mediated by inhaling hot air (11, 15–18). Proposed pathways include activation of bronchopulmonary vagal afferent C fibers, increased cholinergic responses, and stimulation of specific heat shock proteins leading to both epithelial barrier dysfunction and airway inflammation (11, 16, 17).

In addition to advanced age, numerous studies have demonstrated that disproportionate exposure and vulnerability to hazardous heat is influenced by complex interactions between municipal intervention efforts, socioeconomic inequality (19), structural racism (20, 21), access to health care, adaptive capacity, risk perception, and physiological acclimatization and susceptibility (6, 19–23). Urban populations have also been shown to be at an increased risk of heat-related health outcomes due to the urban heat island effect (24) in which the heat retaining and generating properties of the urban form (low albedo surfaces, building configurations that trap heat, diverse sources of heat generation, decreased vegetation, and less evapotranspiration) result in higher air temperatures in urban environments compared to rural areas (25).

In this large, nationwide study of respiratory hospitalizations among Medicare beneficiaries (aged 65 years and older) living in large urban centers, we assessed the role of high ambient temperatures on cause-specific respiratory morbidity between 2000 and 2017. We further quantified the heat-related respiratory burden for specific disease outcomes and assessed whether the burden of all-cause respiratory hospitalizations varied by demographic group and/or geographical setting. A novel contribution of this work defines the relationship between heat and previously unexplored respiratory diagnoses, as well as evaluating the variation in heat-attributable burden and risk across 120 large urban centers.

## METHODS

### Outcome Ascertainment

We obtained short-stay, inpatient hospitalization data for all Medicare beneficiaries aged 65-114 years during 2000-2017 from the Centers for Medicaid and Medicare Services. We excluded hospitalizations for Medicare beneficiaries who were older than 114 years, as well as “long-stay” or “skilled nursing facility” hospitalizations. Medicare records included date of admission, International Classification of Diseases, Ninth and Tenth Revisions diagnosis codes (ICD-9; ICD-10), age, race, sex, indicator of short stay/long stay/skilled nursing facility, and ZIP code of patient residence. Health outcomes of interest were identified using ICD-9 and ICD-10 diagnosis codes in the billing claim (Supplemental Table E1). Five cause-specific respiratory groupings were considered: all-cause, asthma, chronic obstructive pulmonary disease (COPD), respiratory tract infection (RTI), and all other respiratory diseases that did not fit into the previous categories. We refer to this last group as chronic respiratory disease/respiratory failure (CRD/RF) as nearly 90% of CRD/RF hospitalizations were for diagnoses of respiratory failure and progressive inflammatory and fibrotic pulmonary diseases (Supplemental Table E2). Health endpoints were also distinguished based on whether respiratory events were reported in the principal diagnostic code position (principal diagnoses) or the first three diagnostic code positions (first-three diagnoses). Respiratory hospitalizations that occurred within four days of discharge from a prior hospitalization were assumed to be related to the prior event and excluded from the analysis.

Following previous work (24), we included hospitalizations of beneficiaries living in Metropolitan Statistical Areas (MSAs) within the contiguous US with a total 2010 population of 500,000 or more. MSAs with a population greater than 2.5 million were subdivided into metropolitan divisions, as defined by the US Office of Management and Budget, and analyzed as 120 separate study areas, hereafter referred to as large urban centers (LUCs) (Figure E1).

### Ambient meteorological data

Weather station observations of hourly ambient temperature and relative humidity, obtained from the National Oceanic and Atmospheric Administration, were interpolated to census tract centroids using thin-plate spline regressions and aggregated to the daily ZIP code-level (24).

Meteorological variables used in analyses included daily mean (24-hour average), minimum, and maximum temperature, daily mean relative humidity, and daily mean and maximum heat index calculated using the *weathermetrics* package in R. The analysis was restricted to the warm season, defined as June through September, the four warmest months in all large urban centers (LUCs) during the study period (26).

### Statistical Analyses

We applied a two-stage modeling approach to estimate associations between daily ZIP-code level ambient temperature and respiratory hospitalizations within each LUC and nationwide. In the first stage, associations were estimated for each LUC using time-stratified conditional quasi-Poisson regression with distributed lag nonlinear models (27, 28), conditioning on ZIP code of residence, year, month, and day of the week of hospitalization (29). In primary analyses, we defined the exposure-response function with a natural cubic spline and two internal knots placed at the 33rd and 66th percentiles (REFS) of the LUC warm-season temperature distribution based on ZIP code level temperatures within an LUC across all years. The lag-response association was defined using a natural cubic spline with three internal knots equally spaced on the log-scale. A maximum lag window of seven days was chosen *a priori* to account for the effects of the exposure distributed over time (30–35). All models controlled for Federal holidays, same-day (lag day 0) average humidity using a natural cubic spline with four degrees of freedom, and residual temporal trends using natural cubic splines on day of the warm season (four degrees of freedom). Separate models were fitted for each outcome, and for all-cause respiratory outcomes across the following population groups: sex (male, female); age group (65–74, 75–84, 85–114); and race (Black, white).

In the second stage, we pooled the risk estimates using multivariate meta-regression to obtain nationwide effect estimates (36, 37). The meta-regression included LUC average ambient temperature and temperature range (difference between minimum and maximum temperature) as fixed effects to explain variation across LUCs, as done previously (24, 38, 39). We used the best linear unbiased prediction (BLUP) to update the estimates of the LUC-specific associations (24, 36). Sensitivity analyses assessed whether results were robust to model specification (additional details are available in the supplemental material).

For all models, the cumulative effect is the sum of the relative risk (RR) estimates across lag days 0-6 associated with a specific temperature value relative to the LUC-specific warm-season median temperature (reference exposure). We report lag-response associations, cumulative percent change in hospitalizations (%Δ= (RR-1)*100), and 95% confidence intervals (CIs) comparing high warm-season temperatures (95th ambient temperature percentile) to median warm-season temperatures (40–43).

### Attributable Burden Analyses

Attributable burden was quantified at the national, LUC, and subpopulation level using the forward perspective as previously described (44) for the following attributable measures: (1) attributable number (AN), defined as the number of respiratory hospitalizations attributable to temperatures above the 50^th^ percentile; and (2) annual attributable rate (AR), defined as the average annual AN per 100,000 Medicare beneficiaries within subgroup. For the attributable measures, interval uncertainty was obtained empirically through Monte Carlo simulations (44) and are reported as 95% empirical confidence intervals (eCIs). Additional details on the attributable burden calculation are reported in the supplemental material.

## RESULTS

### Descriptive results

In an open cohort of 24.4 million Medicare beneficiaries aged 65-114 living in the 120 largest US LUCs during 2000-2017, we identified 3,275,033 hospitalizations with primary respiratory diagnoses and 8,374,625 hospitalizations with a respiratory diagnosis in the first three diagnostic code positions (Table 1). Pneumonia accounted for over 94% of all respiratory tract infections (Supplemental Table E1). Black beneficiaries comprised the second largest racial/ethnic demographic in the study cohort. Although other race/ethnicities were reported, daily hospitalizations among Asians, Hispanics, and North American Natives were not sufficiently numerous for separate analyses by these race/ethnicities. Cause-specific respiratory hospitalizations had similar proportions by age, sex, and race, except for asthma, which had proportionally more women and Black beneficiaries compared to other outcomes.

**Table 1.** Summary of respiratory hospitalization data, overall and by individual modifying factors, for 120 large urban centers, June-September, 2000-2017.

|  |  | Total hospitalizations, # (%) |  |  |  |  |
| --- | --- | --- | --- | --- | --- | --- |
|  |  | All-cause<br>respiratory | Asthma | COPD | CRD/RF | RTI |
| **Overall** |  |  |  |  |  |  |
|  | Principal dx | 3,275,033 | 161,392 | 763,628 | 1,073,663 | 1,276,350 |
|  | First-three dx | 8,374,625 | 527,677 | 3,076,960 | 3,819,130 | 2,625,698 |
| **Sub-population\*** |  |  |  |  |  |  |
| **Sex** |  |  |  |  |  |  |
|  | Male | 1,438,224 (44%) | 42,077 (26%) | 321,316 (42%) | 499,486 (47%) | 575,345 (45%) |
|  | Female | 1,836,805 (56%) | 119,315 (74%) | 442,312 (58%) | 574,176 (53%) | 701,002 (55%) |
| **Age Group** |  |  |  |  |  |  |
|  | 65-74 | 963,430 (29%) | 61,690 (38%) | 274,411 (36%) | 307,729 (29%) | 319,600 (25%) |
|  | 75-84 | 1,338,840 (41%) | 66,679 (41%) | 339,342 (44%) | 424,600 (39%) | 508,219 (40%) |
|  | 85-114 | 972,763 (30%) | 33,023 (20%) | 149,875 (20%) | 341,334 (32%) | 448,531 (35%) |
| **Race/Ethnicity** |  |  |  |  |  |  |
|  | Black | 351,148 (11%) | 32,824 (20%) | 81,785 (11%) | 117,614 (11%) | 118,925 (9%) |
|  | White | 2,709,189 (83%) | 110,227 (68%) | 641,362 (84%) | 883,374 (82%) | 1,074,226 (84%) |

\*Total number of hospitalizations within the sex, age, and race/ethnicity groups are based on principal causes of the health outcomes. Race/ethnicity percentages do not sum to 100% because all races/ethnicities are not represented in this table. Abbreviations: COPD=Chronic obstructive pulmonary disease; CRD/RF: Chronic respiratory disease/respiratory failure; dx= diagnoses; RTI=Respiratory Tract Infections

Across the study period and all LUCs, warm-season daily mean temperatures ranged from 0.1°C to 42.6°C (Supplemental Table E3). LUCs in the Pacific and Mountain US Divisions experienced the widest warm-season temperature ranges, while temperatures were higher, on average, in LUCs of the South Atlantic and West South Central Divisions (Supplemental Table E3). Across all locations, daily mean warm-season temperatures were highly correlated with other temperature metrics (Supplemental Table E4). Spearman correlations between warm-season mean temperature and warm-season relative humidity within each LUC were more variable and ranged from −0.71 to 0.10 (mean [SD] ρ= −0.28 [0.25], Supplemental Table E4). LUC average warm-season temperature and temperature range helped explain the heterogeneity across locations, as meta-regression I^2^ statistics were <15% for all outcomes and all population subgroups (Supplemental Table E5). LUC-specific daily mean temperature summary statistics are reported in Supplemental Table E6.

### Nationwide Risk and Burden

Nationwide, 7-day cumulative associations at high warm-season temperatures resulted in a 1.2% (95% CI: 0.4, 2.0) increase in hospitalizations for primary diagnoses of all-cause respiratory diseases, primarily driven by increases in RTI [1.8% (95% CI: 0.6, 3.0)] and CRD/RF hospitalizations [1.2% (95% CI: 0.0, 2.4)] (Table 2, Figure 1). For these outcomes, associations monotonically increased with increasing temperatures above the 50^th^ percentile of the warm-season temperature distribution (Figures 1-2; Supplemental Figure E2). We did not observe associations between high warm-season temperatures and asthma or COPD hospitalizations (Figure 1; Supplemental Figure E2). For all outcomes, principal diagnoses had a stronger association with high warm-season temperatures compared to the first-three diagnoses (Figure 1-2; Supplemental Figure E2). Given this finding, only principal diagnosis outcomes were considered in subgroup and LUC specific analyses.

**Figure 1.**
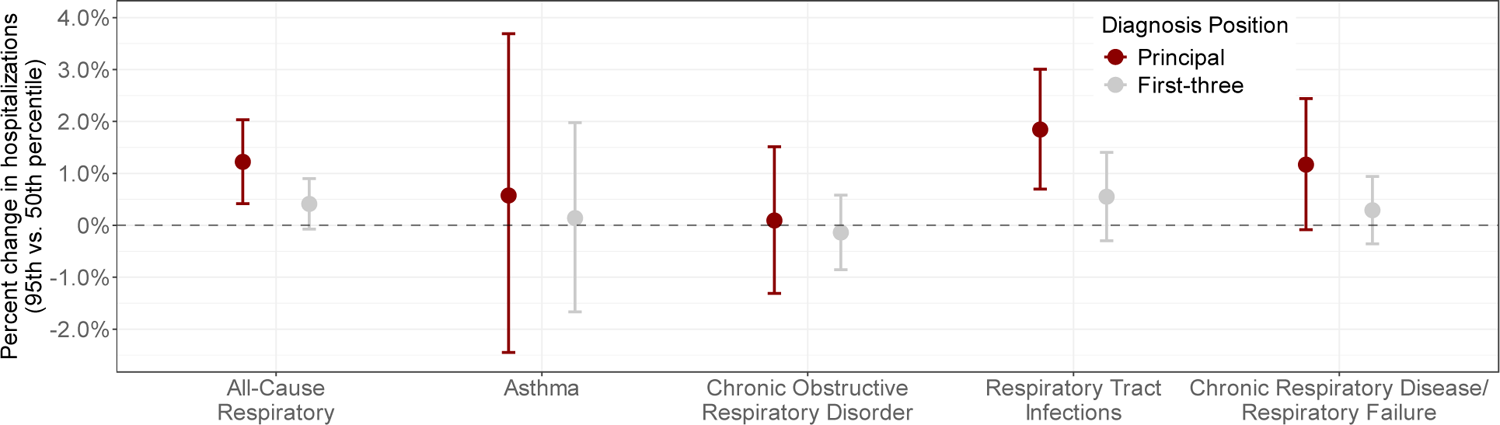
Nationwide 7-day cumulative percent change in hospitalizations and 95% confidence intervals (CIs) between high warm-season temperature and cause-specific respiratory hospitalizations pooled across 120 large urban centers, June-September, 2000-2017. Percent increase in hospitalization compares hospitalizations on days of high warm-season temperature (95^th^ percentile) to median temperature (reference exposure). Percent changes in red represent associations between ambient temperature and respiratory hospitalizations reported in the first diagnostic code position (principal cause of hospitalization), while percent changes reported in grey represent associations between ambient temperature and respiratory hospitalizations reported in the first-three diagnostic code positions.

**Figure 2.**
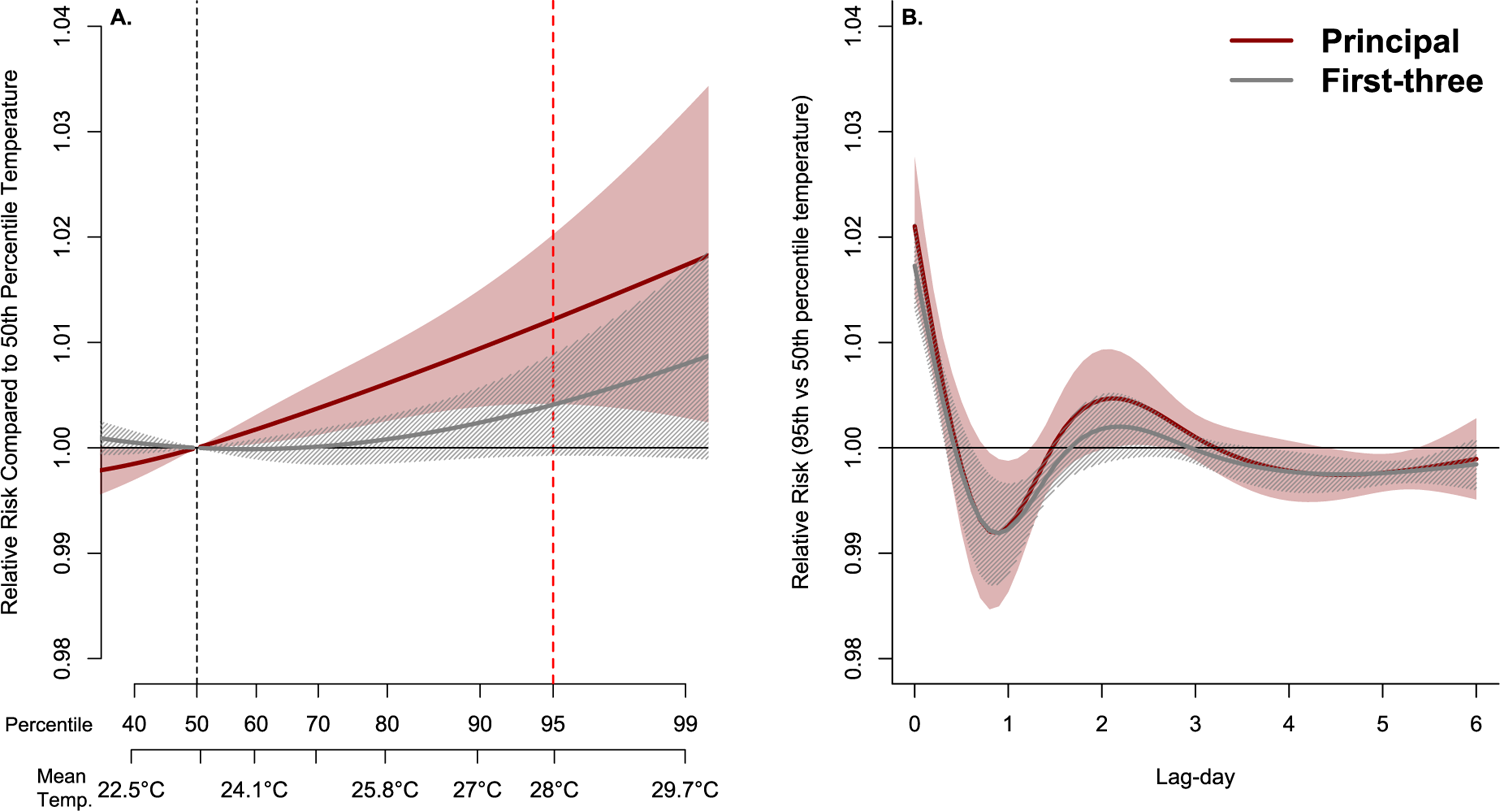
Relative risks (RRs) and 95% confidence intervals (CIs) between temperature and all-cause respiratory hospitalizations pooled across 120 large urban centers, June-September, 2000-2017. Panel A shows the overall 7-day cumulative, nationwide exposure-response relationship between increases in daily average temperature and all-cause respiratory hospitalizations. Panel B demonstrates the nationwide lag-response association for each lag day comparing a day of high warm-season temperature (95^th^ percentile) to median temperature (reference exposure). Associations for principal respiratory causes of hospitalization are reported in red. Associations for respiratory hospitalizations reported in the first three diagnosis codes are reported in grey. Dashed black line indicates the temperature percentile value used as the centering point for temperature contrasts (50^th^ percentile). The dotted red line indicates the 95^th^ percentile (high warm-season temperature). Color bands around solid lines represent 95% CIs. Abbreviations: CI= confidence intervals; RR: relative risk; Mean temp. = daily mean temperature

**Table 2.**
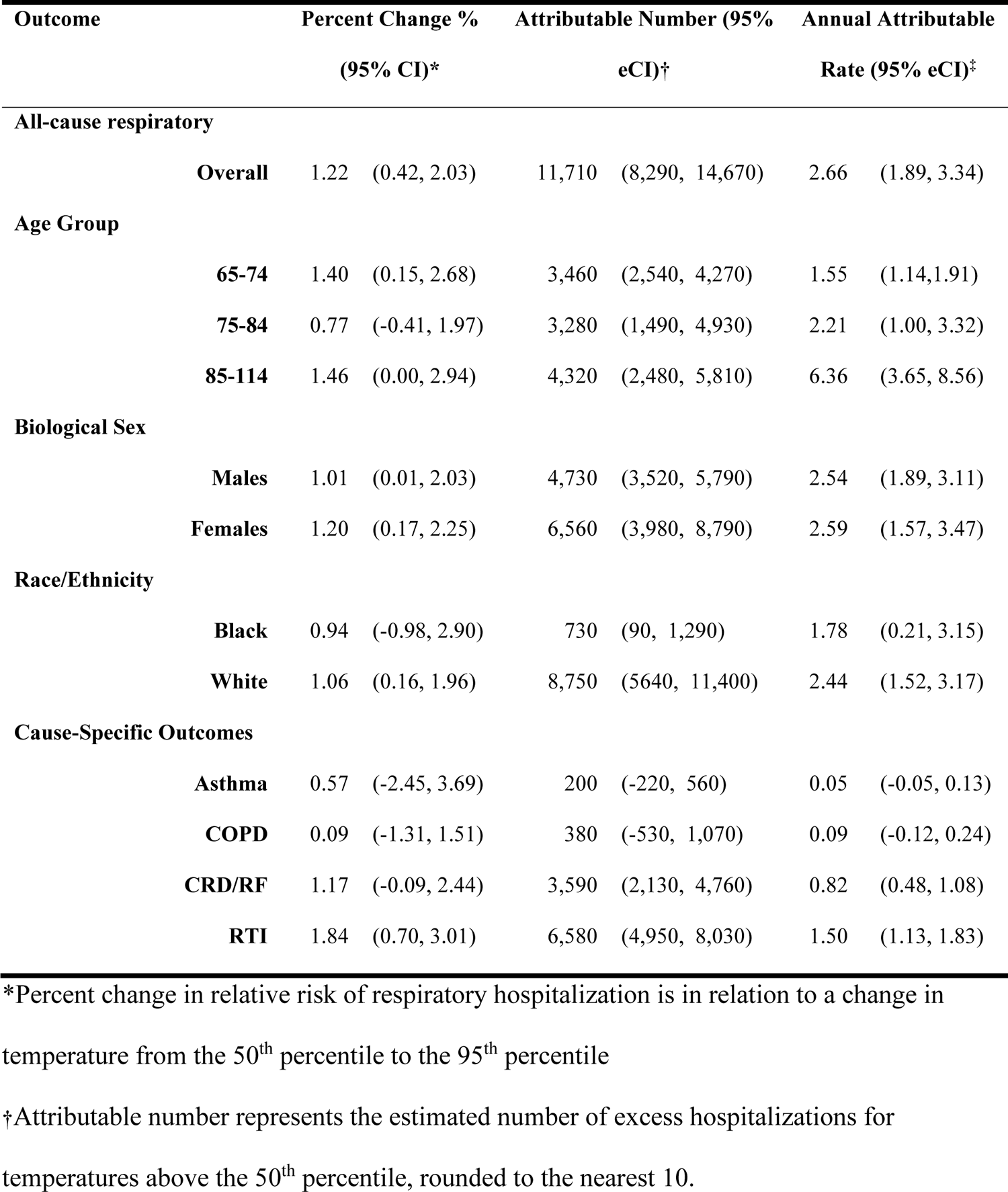
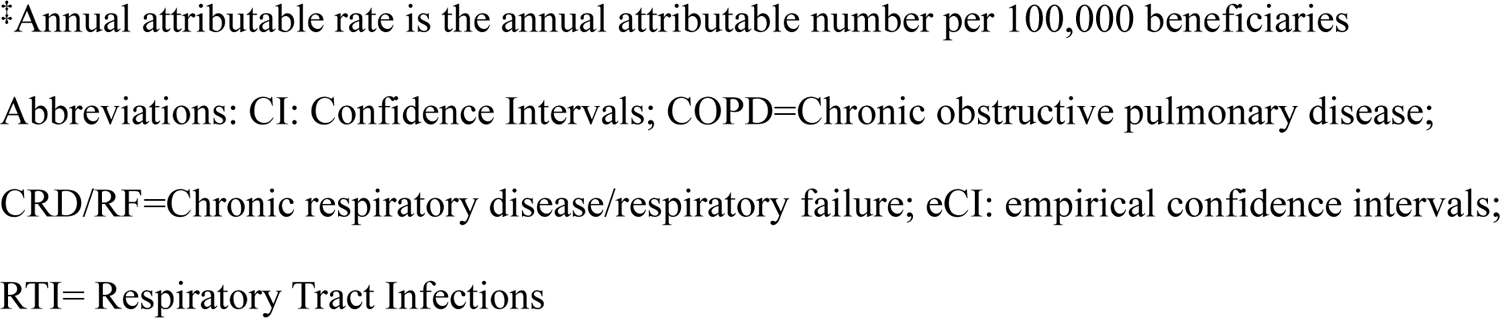
Seven-day cumulative percent change in relative risks, attributable number, and annual attributable rates between all-cause respiratory hospitalizations and high warm-season temperature for 120 large urban centers, June-September, 2000-2017.

| Outcome | Percent Change % |  |  | Attributable Number (95% |  | Annual Attributable |  |
| --- | --- | --- | --- | --- | --- | --- | --- |
|  |  | (95% CI)* |  | eCI)† |  | Rate (95% eCI)‡ |  |
| All-cause respiratory |  |  |  |  |  |  |  |
|  | Overall | 1.22 | (0.42, 2.03) | 11,710 | (8,290, 14,670) | 2.66 | (1.89, 3.34) |
| Age Group |  |  |  |  |  |  |  |
|  | 65-74 | 1.40 | (0.15, 2.68) | 3,460 | (2,540, 4,270) | 1.55 | (1.14,1.91) |
|  | 75-84 | 0.77 | (-0.41, 1.97) | 3,280 | (1,490, 4,930) | 2.21 | (1.00, 3.32) |
|  | 85-114 | 1.46 | (0.00, 2.94) | 4,320 | (2,480, 5,810) | 6.36 | (3.65, 8.56) |
| Biological Sex |  |  |  |  |  |  |  |
|  | Males | 1.01 | (0.01, 2.03) | 4,730 | (3,520, 5,790) | 2.54 | (1.89, 3.11) |
|  | Females | 1.20 | (0.17, 2.25) | 6,560 | (3,980, 8,790) | 2.59 | (1.57, 3.47) |
| Race/Ethnicity |  |  |  |  |  |  |  |
|  | Black | 0.94 | (-0.98, 2.90) | 730 | (90, 1,290) | 1.78 | (0.21, 3.15) |
|  | White | 1.06 | (0.16, 1.96) | 8,750 | (5640, 11,400) | 2.44 | (1.52, 3.17) |
| Cause-Specific Outcomes |  |  |  |  |  |  |  |
|  | Asthma | 0.57 | (-2.45, 3.69) | 200 | (-220, 560) | 0.05 | (-0.05, 0.13) |
|  | COPD | 0.09 | (-1.31, 1.51) | 380 | (-530, 1,070) | 0.09 | (-0.12, 0.24) |
|  | CRD/RF | 1.17 | (-0.09, 2.44) | 3,590 | (2,130, 4,760) | 0.82 | (0.48, 1.08) |
|  | RTI | 1.84 | (0.70, 3.01) | 6,580 | (4,950, 8,030) | 1.50 | (1.13, 1.83) |

\*Percent change in relative risk of respiratory hospitalization is in relation to a change in temperature from the 50<sup>th</sup> percentile to the 95<sup>th</sup> percentile
†Attributable number represents the estimated number of excess hospitalizations for temperatures above the 50<sup>th</sup> percentile, rounded to the nearest 10.
‡Annual attributable rate is the annual attributable number per 100,000 beneficiaries
Abbreviations: CI: Confidence Intervals; COPD=Chronic obstructive pulmonary disease;
CRD/RF=Chronic respiratory disease/respiratory failure; eCI: empirical confidence intervals;
RTI= Respiratory Tract Infections

For all-cause respiratory diseases, RTIs, COPD, and CRD/RF, the pooled lag-response associations demonstrated a robust increase in risk on lag day 0, followed by a decline in risk on lag day 1 possibly due to morbidity displacement, and weak or slightly negative risk across later lag periods (Figure 2; Supplemental Figure E2). For all-cause respiratory hospitalizations, we observed a 2.1% (1.5, 2.8) increase in hospitalizations on lag day 0, with attenuation on the following 6 days (Figure 2). Across all study locations and years, we estimated 11,710 (95% eCI: 8,290-14,670) excess all-cause respiratory hospitalizations due to warm-season temperatures above the 50^th^ percentile (Table 2). Results from *Sensitivity Analyses* are reported in the supplemental material.

### Subgroup-Specific Risk and Burden

We did not observe effect measure modification in relative risk (RR) by age group, sex, or race/ethnicity at the national level (Table 2; Supplemental Figure E5). Due to very few observed hospitalizations among Black beneficiaries, we were unable to estimate associations for three LUCs (Provo-Orem, UT; Boise City, ID; and Portland-South Portland, ME). The burden of heat-related respiratory hospitalizations was largest among beneficiaries aged 85 and over, as reflected in the ARs (Table 2). Attributable burden at the national-level was similar among males and females (Table 2).

### LUC-Specific Risk and Burden

We observed considerable geographic variation in the magnitude and direction of heat-related relative risk (RR) and attributable risk (AR) across the study areas. For heat-related all cause respiratory diseases, percent change in hospitalizations ranged from a 3.3% decrease to a 4.7% increase, and ARs ranged from −7.6 to 11.9 annual excess hospitalizations per 100,000 beneficiaries (Figure 3). Across the nine US Census divisions, the highest attributable burden rates were observed in LUCs located in the Pacific, East North Central (a subregional division of the Midwest), New England, and Mid-Atlantic (Figure 3; Supplemental Figure E6; Supplemental Table E8). Although the 58 LUCs in these divisions only accounted for 56% (1,826,777) of the total number of respiratory hospitalizations, 98.5% (11,539) of all heat-related excess respiratory hospitalizations between 2000-2017 were reported in these LUCs. The lowest RRs and ARs were observed in LUCs in the South Atlantic region, especially Florida (Figure 3; Supplemental Figure E6; Supplemental Table E8). RTI and CRD/RF accounted for most of the heat-related respiratory hospitalization burden across all LUCs (Supplemental Figure E7).

**Figure 3.**
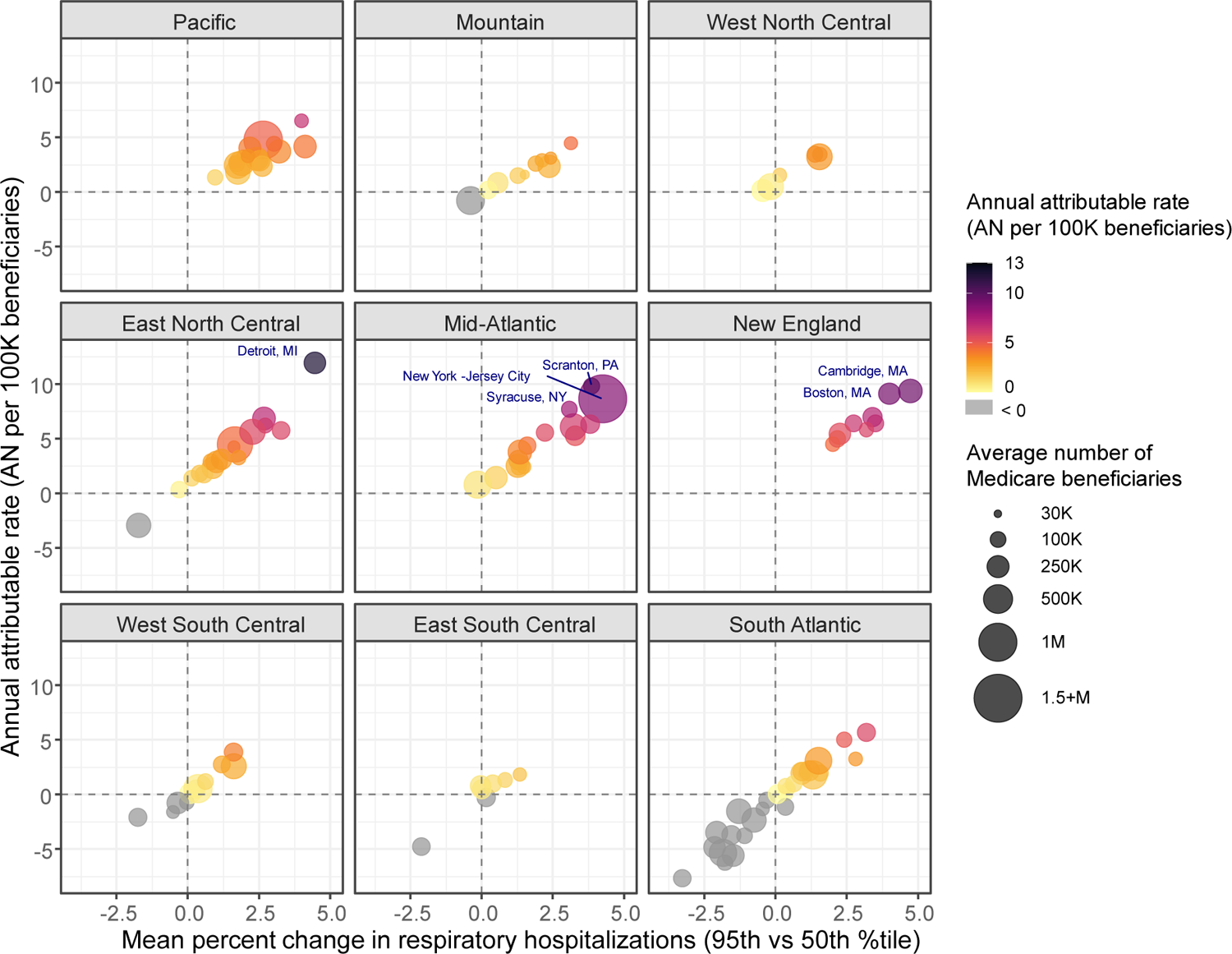
Percent change and attributable burden of high temperature-related all-cause respiratory hospitalizations for 120 large urban centers (LUCs) by US Division, June-September, 2000-2017. LUC-specific percent changes in respiratory hospitalizations are shown by LUC-specific annual attributable number of excess hospitalizations per 100,000 beneficiaries (annual attributable rate) for each US division. For each location, the color of the circle is determined by LUC-specific annual attributable rate, and the size of the circle is determined by the number of Medicare beneficiaries within each LUC. Annotated study area names represent LUCs (n=6) in the top 5% of annual attributable rates, where Medicare beneficiaries experience the greatest burden of heat-related respiratory morbidity. AN=Attributable Number; LUC=large urban center

At the metropolitan-level we observed a disproportionate burden of heat-related all-cause respiratory hospitalization across age groups, sex, and race. In contrast to the overall population, the highest burden among Black beneficiaries was observed in LUCs of the South Atlantic and East South-Central divisions (Figure 4; Supplemental Figure E8). In the mid-Atlantic and Pacific divisions, the burden was more evenly distributed among white and Black beneficiaries. In the Mountain, West North Central, and New England divisions - which have much fewer Black beneficiaries – the white population accounted for most of the burden (Supplemental Figure E8). Across age groups, the 85 and over population accounted for most of the burden in almost every metropolitan area (Supplemental Figure E9). On average, burden rates were similar between males and females across most US divisions. However, we consistently observed higher ARs among males living in LUCs of the Mountain and West North Central divisions (Supplemental Figure E10).

**Figure 4.**
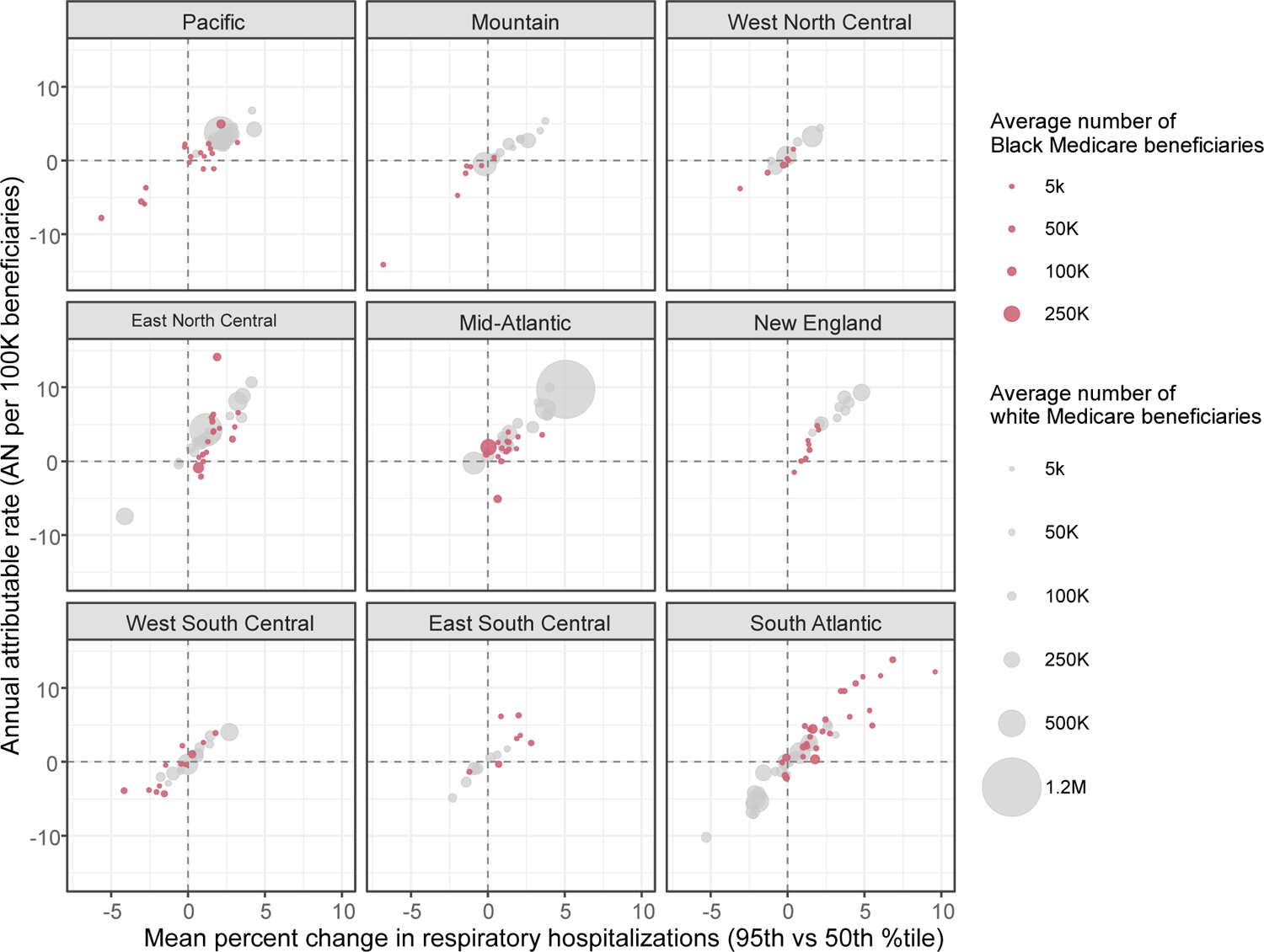
Percent change and attributable burden of heat-related all-cause respiratory hospitalizations among white and Black beneficiaries for 120 large urban centers (LUCs) by US Division, June-September, 2000-2017. LUC-specific percent changes in respiratory hospitalizations are shown by LUC-specific annual attributable number of excess hospitalizations per 100,000 beneficiaries (annual attributable rate) for Black and white beneficiaries for each US division. Red circles represent risk estimates among Black beneficiaries and grey circles represent risk estimates among white beneficiaries. The size of the circle is determined by the number of Medicare beneficiaries within each study location. AN=Attributable Number; LUC=large urban center

## DISCUSSION

In this nationwide study, we characterized the relationship between short-term exposure to high warm-season temperature and cause-specific respiratory morbidity among older populations living in the 120 largest US urban centers. Notably, we observed that elevated ambient temperature led to excess hospitalizations for respiratory failure and chronic inflammatory and fibrotic diseases such as pneumonitis, pleural effusion, and interstitial pulmonary diseases.

Across demographic groups, most of the burden was attributed to the oldest age group (85+), and most heat-related excess hospitalizations were due to exacerbation of RTI and CRD/RF. Finally, demographic and LUC results suggest heat-related health disparities were likely driven by complex social and contextual forces acting on individuals to intensify their exposure and respiratory health risk.

To our knowledge, this study is the first large population-based study in the US to define a relationship between ambient high temperature and hospitalizations for respiratory failure and chronic inflammatory and fibrotic respiratory diseases (pneumonitis, pleural effusion, and interstitial pulmonary diseases). A nascent literature base on the biological effects of high ambient temperature on respiratory injury points to a few key mechanisms involving inflammatory responses and epithelial barrier permeability that may act independently or in concert to initiate and sustain pulmonary injury (11, 16, 17, 45, 46). Possible mechanisms being investigated include activation of specific heat-shock proteins, which have been associated with respiratory disease progression of both asthma and fibrosis and may function through dysregulation of tight junction permeability. However, so few mechanistic studies have been performed that the collective biological effects of hot, humid air on the respiratory tract are still to be discovered.

The results of this study underscore the importance of considering geographic and socio-demographic factors when assessing heat-health risks. At the national level we did not observe effect modification by sex, age, or race; however, LUC and regional analyses suggest a disproportionate burden of heat-related respiratory hospitalizations among Black beneficiaries compared to their white counterparts across LUCs in the South Atlantic and East South Central Divisions. In these locations, it is possible that structural racism manifests differently than in other parts of the country and intersects with physiological and contextual factors (e.g., comorbidities and cumulative impacts of environmental and social stressors) to disproportionately burden Black beneficiaries. The increasingly warming climate is likely to deepen existing disparities resulting from past and present structural racism. We also observed higher heat-related excess hospitalization rates among beneficiaries of advanced age (85 years and older) compared to other age groups in most LUCs. While age-related susceptibility to extreme heat is well established, this study is among the first to demonstrate the disproportionate burden of heat on the respiratory health of older adults across nearly every major US metropolitan area.

In the overall Medicare population, risks were generally lowest in the Southeast and Southwest, possibly due to higher prevalences of air conditioning (47, 48) and greater physiological adaptation to heat. Findings of lower heat-related mortality and morbidity risk in communities with hotter summers compared to milder summers are commonly reported in multi-city, multi-national studies (4, 15, 43). However, the southwestern and southeastern US geographies are expected to experience more severe extreme heat over the coming decades and human heat acclimatization capacity has a physiological ceiling that may not be able to overcome projected temperature increases without behavioral and technological adaptations (12, 13). This study also identified LUCs in the Pacific, the East North Central, New England, and the Mid-Atlantic that are particularly vulnerable to the effects of extreme heat exposure, with high risk and high burden rates among the Medicare population (Figure 3). On average, LUCs in these US divisions have historically lower prevalence of central air conditioning (47, 48), mild summers, and a wide range of warm-season temperature (Supplemental Table E4, Supplemental Table E6). In addition, a recent study suggests that high temperature warning thresholds may be too high in colder US locations and may not take into consideration the sensitivity of the local population (49).

Overall, the nationwide pooled risk estimates for all-cause respiratory disease were in the range of risk estimates reported in other heat-hospitalization studies among Medicare beneficiaries. In this study, we observed a 1.22% increase in all-cause respiratory hospitalizations across lag days 0-6. Other Medicare based studies, with similar cumulative lag considerations and outcome definitions, report heat-related increases in all-cause respiratory hospitalizations ranging from 0.0-4.0% for a change in the previous week’s ambient heat (6, 15). In contrast to previous studies, we did not detect a cumulative association between heat and COPD. However, we observed elevated COPD exacerbation risk on lag day 0 [1.70 (1.04, 2.36)] with a robust negative association possibly due to morbidity displacement on lag day 1 [−1.47 (−2.12, −0.80)], and no association across later lag periods. Differences in study locations, study-period, heat exposure metrics, the functional form of temperature, and how we defined a health relevant change in temperature (95th percentile to median temperature) could explain differences in reported risk estimates.

The ability to evaluate heat-related respiratory hospitalizations among older adults across 120 LUCs was facilitated by an 18-year study period with rich patient level data and fine-scale meteorological data. However, additional considerations should be acknowledged when interpreting results. First, this study is not able to account for air conditioning prevalence and other potentially important housing characteristics due to limited data availability. Populations that do not have access to air conditioning or cannot afford to adequately cool their homes likely experience higher levels of heat exposure and could be at greater risk. As other studies have indicated, heat-related mortality and morbidity risk may be in competition in some locations.

Thus, high rates of heat-related mortality could artificially create a protective effect when examining heat-related hospitalizations (6, 8, 50). Given the use of spatially interpolated weather station data aggregated to the ZIP code level, there could be exposure misclassification error that would likely result in attenuation toward the null. Finally, reported risks for the entire Medicare population are largely driven by the strength of the association among white beneficiaries living in large LUCs (Supplemental Figure E10) and may not be generalizable to other races/ethnicities or to rural populations.

## CONCLUSION

This study considerably extends the current understanding of the relationship between heat and respiratory hospitalizations among older adults by characterizing novel respiratory endpoints that have not been previously examined in a large, multi-city epidemiologic study. The findings raise new questions on the role that heat may play in relation to respiratory failure and airway diseases like interstitial lung disease, pneumonitis, and pleural effusion. Additionally, we observed disparities in the impact of heat on the respiratory health of Black beneficiaries that were geographically dependent. These results suggest that the drivers of heat-related respiratory morbidity are likely due to the joint effects of physiological susceptibility and contextual forces acting on individuals to intensify their exposure and risk. Additional studies to further explore these disparities, as well as the effects of heat on understudied airway diseases, are needed to corroborate our findings. Mechanistic studies, especially controlled exposures, will be essential to determine whether the associations identified in this study are causative in nature. Findings from this study and similar ones can be used at the local level to motivate intervention studies/efforts and repeated studies can aid in monitoring progress towards reducing heat-related health disparities.

## Data Availability

Medicare hospitalization data are restricted by IRB protocols and data use agreements with Centers for Medicaid and Medicare (CMS) but other researchers may obtain the same Medicare data directly from CMS. Daily meteorological data are publicly available from the National Oceanic and Atmospheric Administrations National Climatic Data Centers Global Surface Summary of the Day database. American Community Survey (ACS) data are publicly available through the Census Bureau.

## Acknowledgements

The authors would like to acknowledge the important contributions of members of Clinical Research Branch (Wei-Lun Tsai, Corinna Keeler, Kathryn Burns, Riley Short, William Steinhardt, Cavin Ward-Caviness) of the US EPA who have facilitated our ability to examine climate influenced exposures on health. The successes of this manuscript and its future impacts are largely due to the hard work of behind-the-scenes researchers whose combined contribution to this study has been invaluable.

## Supplemental Material

### ADDITIONAL DETAIL OF THE METHODS

#### Study Areas

To investigate the health effects of extreme heat on urban populations, this study included the 120 largest US metropolitan areas (the names of each metropolitan area are reported in Table E6). Metropolitan areas were identified using US Office of Management and Budget definitions for Metropolitan Statistical Areas (MSA). Similar to Cleland et al., 2023(E1), we used the 2010 Decennial Census survey to restrict our analysis to MSAs within the contiguous US with a total 2010 population of 500,000 or more. Metropolitan Statistical Areas with a population greater than 2.5 million were subdivided into metropolitan divisions, according to OMB definitions, and analyzed as separate study areas. For example, the metropolitan divisions, Anaheim-Santa Ana-Irvine, CA and Los Angeles-Long Beach-Glendale, CA are considered two separate study areas in our analyses; however, these metropolitan divisions make up the Los Angeles-Long Beach-Anaheim, CA MSA. We refer to our 120 study areas as Large Urban Centers (LUCs). In our analyses, all residential ZIP codes that fully or partially overlapped with Metropolitan Statistical Area boundaries were included. ZIP codes were excluded if they were classified as Post Office Boxes, businesses, or universities. The resulting study included 11,924 ZIP codes. For regional analyses, large urban centers (LUCs) were grouped according to the nine US Census divisions.

### Additional methodological details

#### Extracting hospitalization records

We extracted patient records and summary files from Medicare billing claims data for short stay, in-patient hospitalizations for any respiratory event reported in the first 3 diagnostic code positions from 2000-2017. Respiratory events were identified using International Classification of Diseases, Ninth and Tenth Revisions codes, specifically, 460-519 (ICD-9) and J00-J99 (ICD-10). We extracted data only for the 11,924 ZIP codes included in our study locations.

#### Outcomes

“Principal” respiratory hospitalizations were explored based on ICD codes reported in the principal diagnosis code position. “First-three” respiratory hospitalizations were explored based on ICD codes reported in the first three diagnosis code positions. We created several cause-specific outcomes using ICD codes (Table E1).

#### Warm-Season

We defined the warm-season as June-September as these were the four warmest months on average from 2000-2017 in each study location. To account for N/A values when creating the 7-day lags, we utilized hospitalization data from May 25 to September 30^th^.

#### Reference exposure

The LUC-specific median temperature was used as the reference exposure. We defined a health relevant change in temperature as a change from the 50^th^ to 95^th^ percentile of the warm-season temperature distribution. In this study, we used median temperature as the reference exposure instead of the minimum hospitalization percentile to facilitate risk communication and to allow for easier comparisons across outcomes, demographic groups, and alternative exposure metrics as the minimum hospitalization percentile varied across these factors (Table E3).

### Statistical Analyses

#### Stage 1 – Large Urban Area specific models

In the first stage of our statistical approach we implemented time-stratified conditional quasi-Poisson regression for each location matching on day, month, year, and ZIP code of Medicare beneficiary residence

- Conditional Poisson modeling was performed for each study location for each outcome and for each subgroup
- Non-linear temperature exposure-response associations were explored by placing 2 internal knots at 33^rd^ and 66^th^ percentiles of the warm-season temperature distribution
- Cumulative effects were explored across lag days 0-6 using DLNM with 3 internal knots equally spaced on the log scale
- We specified quasi-Poisson (family=”quasipoisson”)
- We excluded empty strata (subset=keep)
- We excluded missing data (na.action = “na.exclude”)

The general form of the Stage 1 model for each study location is as follows:

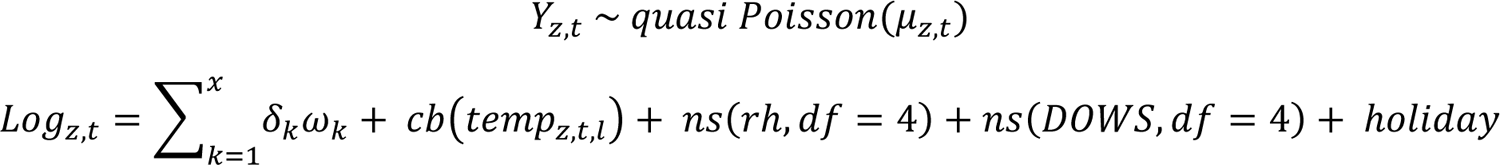

where, Yzt indicates the daily number of hospitalizations in ZIP code z on day t, and t indexes the event (hospitalization) and control days. ωk denotes the indicator variables that distinguish the case–control sets for each ZIP code, x is the total number of case–control sets (11,924), and δk denotes parameters specific to the case–control sets (which are not estimated in conditional Poisson regression). cb(tempz,t,l) is the crossbasis function for daily ambient temperature in ZIP code z at time t and lag l (in this case 7 days). Other model covariates included a natural cubic spline with 4 degrees of freedom for same day relative humidity (rh), a natural cubic spline with 4 degrees of freedom for day of the warm-season (DOWS), and a binary indicator variable for Federal holidays.

#### Stage 2 - Multivariate meta regression pooling 120 LUC-specific estimates

In the second stage of our statistical approach, estimates from LUC-specific stage 1 models were reduced to the cumulative risk during the lag period for each location.

- We pooled 120 LUC-specific estimates using multivariate meta-regression with fixed effects for LUC-specific average warm-season temperature and LUC-specific temperature range to account for possible effect modification on the pooled relationship.
- For all models, the cumulative effect is the sum of the relative risk (RR) estimates across lag days 0-6 associated with a specific temperature value relative to the LUC-specific warm-season median temperature (reference exposure). We report lag-response associations, cumulative percent change in hospitalizations (%Δ= (RR-1)*100), and 95% confidence intervals (CIs) comparing high warm-season temperatures (95th ambient temperature percentile) to median warm-season temperatures.
- We report the I^2^ heterogeneity statistic for all meta-regressions (Table E5).
- Best Linear Unbiased Predictions (BLUP) were extracted for each city and used to report LUC-specific RRs and 95% CIs that were “shrunk” based on the underlying distribution across all 120 study locations.

#### Attributable burden and excess hospitalizations

- BLUPs were used to estimate the number of excess hospitalizations (attributable number) within each ZIP code over the next 7 days for each day of the series above the ZIP code-specific 50^th^ percentile of the warm-season distribution. Attributable numbers were summed across all days and ZIP codes to derive LUC-level burdens for all outcomes and subgroups, as well as a nationwide burden estimate, for all outcomes and subgroups.
- Specifically, the following equation was used:

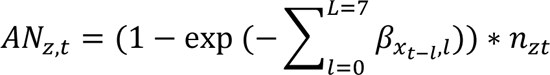

Where *n* is the total number of respiratory hospitalizations in ZIP code *z* on day *t*. β_xt−1,l_ represents the LUC-specific log relative risk (based on BLUP estimates) between ambient temperature (*x*_*t*−1_) and respiratory hospitalizations at lag *l*. The attributable burden calculation was restricted to days when temperatures were above the 50^th^ percentile of the warm-season temperature distribution for each ZIP code. Empirical 95% CIs for the burden estimates were obtained through Monte Carlo simulations, using 1,000 iterations and assuming a multivariate normal distribution of the location-specific BLUP of reduced coefficients (E2). To estimate the excess heat-related respiratory hospitalizations, for each metropolitan area we summed *AN*_*zt*_ and the Monte Carlo estimates across all days and ZIP codes within each metropolitan area. To derive the nationwide estimate, we summed *AN*_*zt*_ and the Monte Carlo estimates across all days and all ZIP codes.

- The total attributable number for each LUC and all 120 LUCs was divided by 18 and by the annual average number of beneficiaries in each LUC and across all locations to estimate an *annual* attributable rate that is adjusted for the Medicare beneficiary population. *Annual* attributable rates are expressed as the average annual number of excess hospitalizations per 100,000 beneficiaries overall and within each subgroup.
- We calculated empirical confidence intervals (eCI) using Monte Carlo simulations assuming a multivariate normal distribution of the BLUP of reduced coefficients.

### Sensitivity Analyses - Approach

#### Sensitivity analyses were performed for three purposes

1. To determine appropriate temporal and confounder control;
2. To evaluate whether results were robust to model specification; and
3. To evaluate whether results and interpretations were consistent across different exposure metrics.

To determine appropriate temporal and time-varying confounder control, we evaluated whether inclusion of time splines during the warm season improved the model fit. We also explored the impact of inclusion of an interaction between year and day of the warm season (used to capture between-year differences in previous studies) (E3, E4). Finally we evaluated model performance using humidity with lagged effects verses same-day humidity.

To evaluate whether results were robust to model specification, we examined knot placement in the exposure-response function comparing models with two evenly spaced internal knots to models with one internal knot placed at either the 50^th^ or 75^th^ percentile of the warm-season temperature distribution, or two internal knots placed at the 50^th^ and 90^th^ percentile. Sensitivity analyses did not explore three internally placed knots. We also estimated associations using unconstrained lag specifications, and constrained lagged effects with two and three internally placed knots.

Finally, we estimated associations across four additional exposure metrics (minimum temperature, maximum temperature, mean heat index, maximum heat index) and lag days 0-3 instead of 0-6 to further explore the sensitivity of our model. When estimating associations between respiratory disease and heat index metrics, we did not include additional control for relative humidity since it was used in the calculation of the heat index. Results of the sensitivity analyses are reported in Supplemental Table E7.

### Sensitivity Analyses - Results

Sensitivity analyses demonstrated that results and interpretations from main analyses were robust to changes in knot placement and lag structure (Table E7). Risk estimates were attenuated when defining heat exposure using minimum and maximum ambient temperature and maximum heat index (Figures E3-4; Table E7). Associations were stronger when using mean heat index as the exposure (%Δ: 1.49% [95%CIs: 0.76, 2.23]) and when estimating associations across lag days 0-3 ([%Δ: 1.54% (95% CIs: 0.86%, 2.23%) compared to lag days 0-6 (Figures E3-4; Table E7).

---

**Figure E1.**
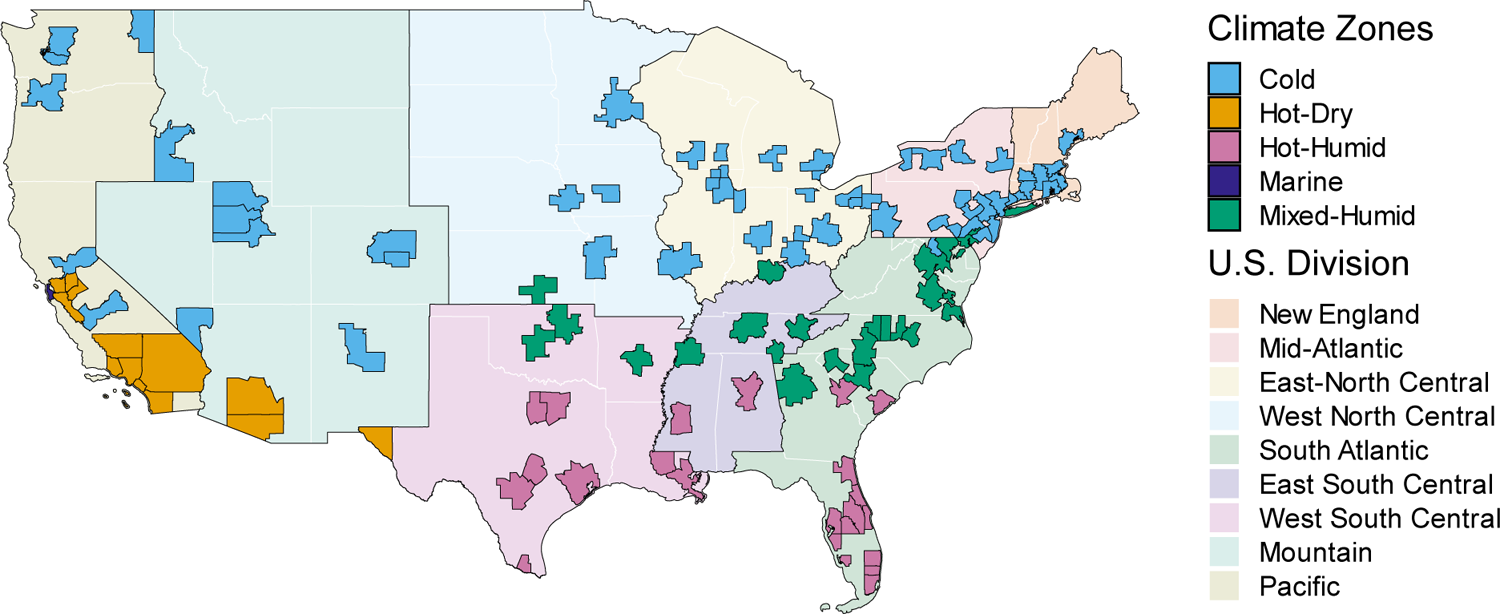
Spatial representation of metropolitan areas included in this study. Metropolitan areas are shown by US Division, and the color scale of the metropolitan areas indicates their IECC climate zones.

**Figure E2.**
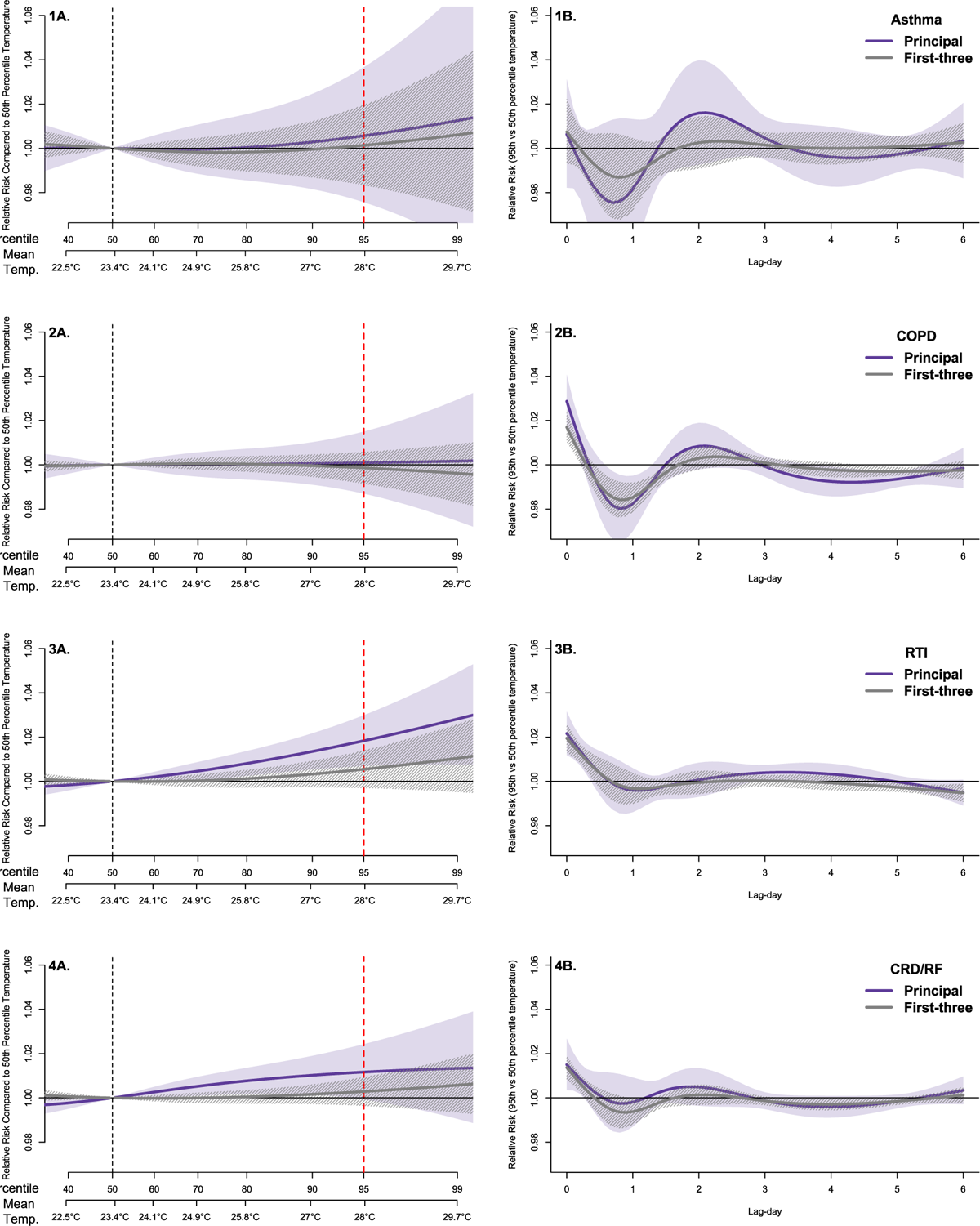
Relative risks (RR) and 95% CIs between ambient mean temperature and cause-specific hospitalizations pooled for 120 metropolitan areas, June-September, 2000-2017. Panels 1A, 2A, 3A, and 4A show the overall 7-day cumulative, pooled exposure-response relationship between increases in daily average temperature and hospitalizations for asthma (1A), COPD (2A), RTI (3A), and CRD/RF (4A). Panels 1B, 2B, 3B, and 4B demonstrate the pooled lag-response association for each lag day comparing a day of high warm-season temperature (95^th^ percentile) to median temperature (reference exposure). Associations for principal diagnoses of cause-specific respiratory hospitalizations are reported in purple. Associations for cause-specific respiratory hospitalizations in the first-three diagnoses are reported in grey. Dashed black line indicates the temperature percentile value used as the centering point for temperature contrasts (50^th^ percentile). The dotted red line indicates the 95^th^ percentile (high warm-season temperature). Color bands around solid lines represent the 95% CIs.

**Figure E3.**
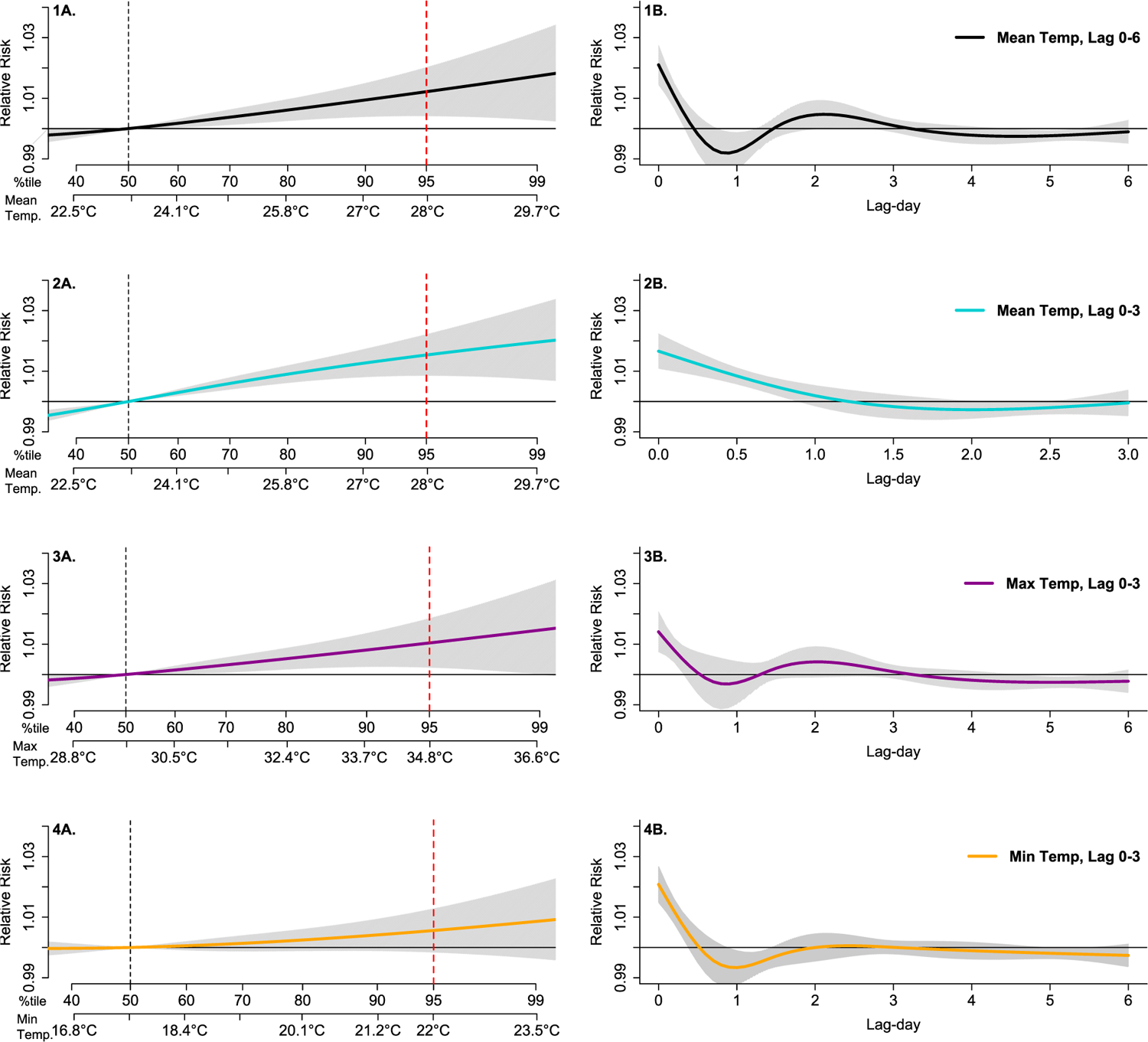
Comparing the impact of using alternative ambient temperature exposure metrics versus mean temperature on cumulative pooled risk estimates. Panels 1A, 2A, 3A, and 4A show the overall 7-day cumulative, pooled exposure-response relationship between increases in all-cause respiratory hospitalizations and the following exposure metrics: (1A) daily mean temperature lags 0-6; (2A) daily mean temperature lags 0-3; (3A) daily maximum temperature lags 0-6; and (4A) daily minimum temperature lags 0-6. Panels 1B, 2B, 3B, and 4B demonstrate the pooled lag-response association for each lag day comparing a day of high warm-season temperature (95^th^ percentile) to median temperature (reference exposure). Dashed black line indicates the temperature percentile value used as the centering point for temperature contrasts. The dotted red line indicates the 95th percentile (extreme heat).

**Figure E4.**
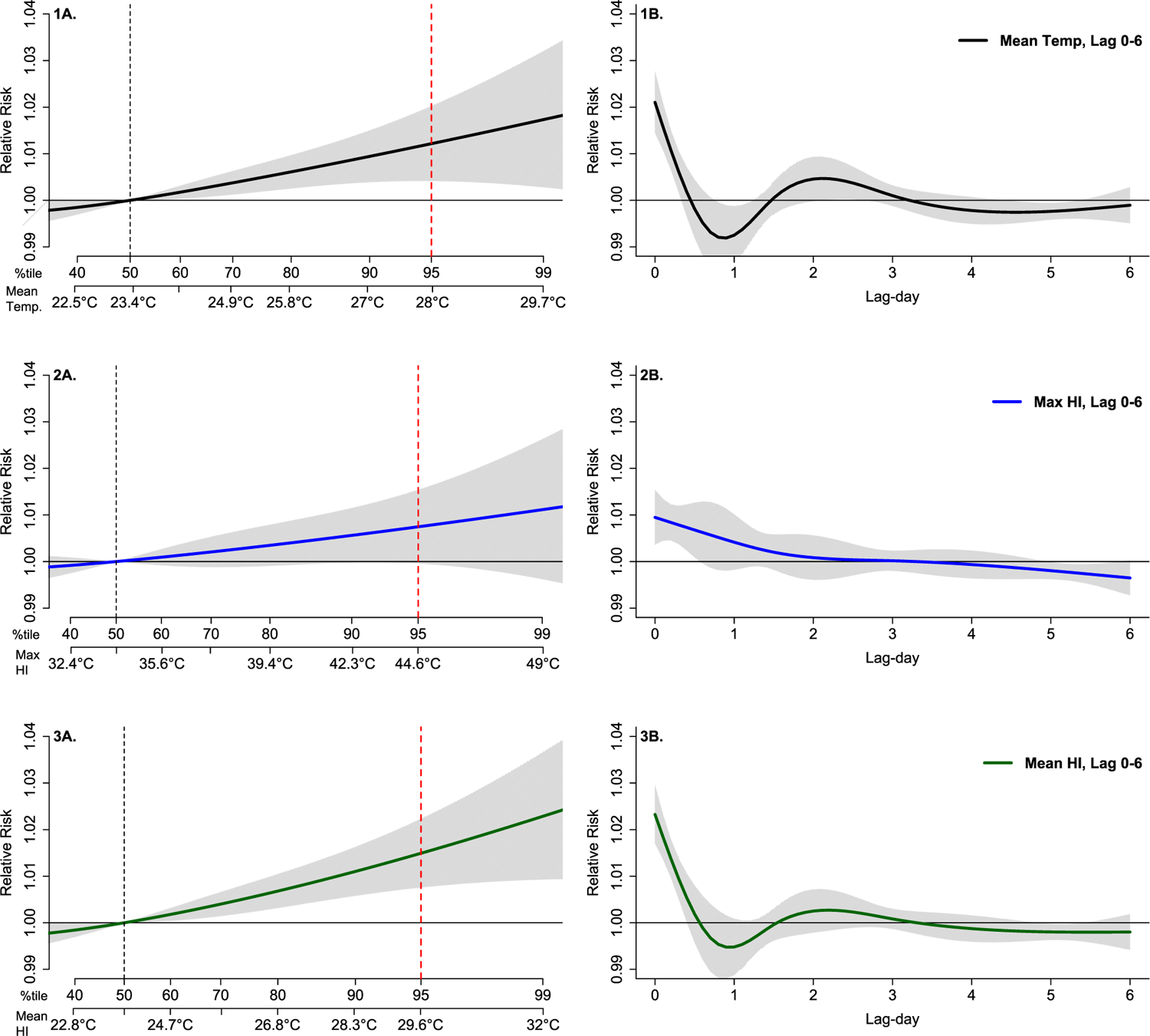
Comparing the impact of using heat index exposure metrics versus mean temperature on cumulative pooled risk estimates. Panels 1A, 2A, 3A show the overall 7-day cumulative, pooled exposure-response relationship between increases in all-cause respiratory hospitalizations and the following exposure metrics: (1A) daily mean temperature lags 0-6; (2A) daily maximum heat index lags 0-6; (3A) daily mean heat index lags 0-6. Panels 1B, 2B, and 3B demonstrate the pooled lag-response association for each lag day comparing a day of high warm-season temperature (95^th^ percentile) to median temperature (reference exposure). Dashed black line indicates the temperature percentile value used as the centering point for temperature contrasts. The dotted red line indicates the 95th percentile (extreme heat).

**Figure E5.**
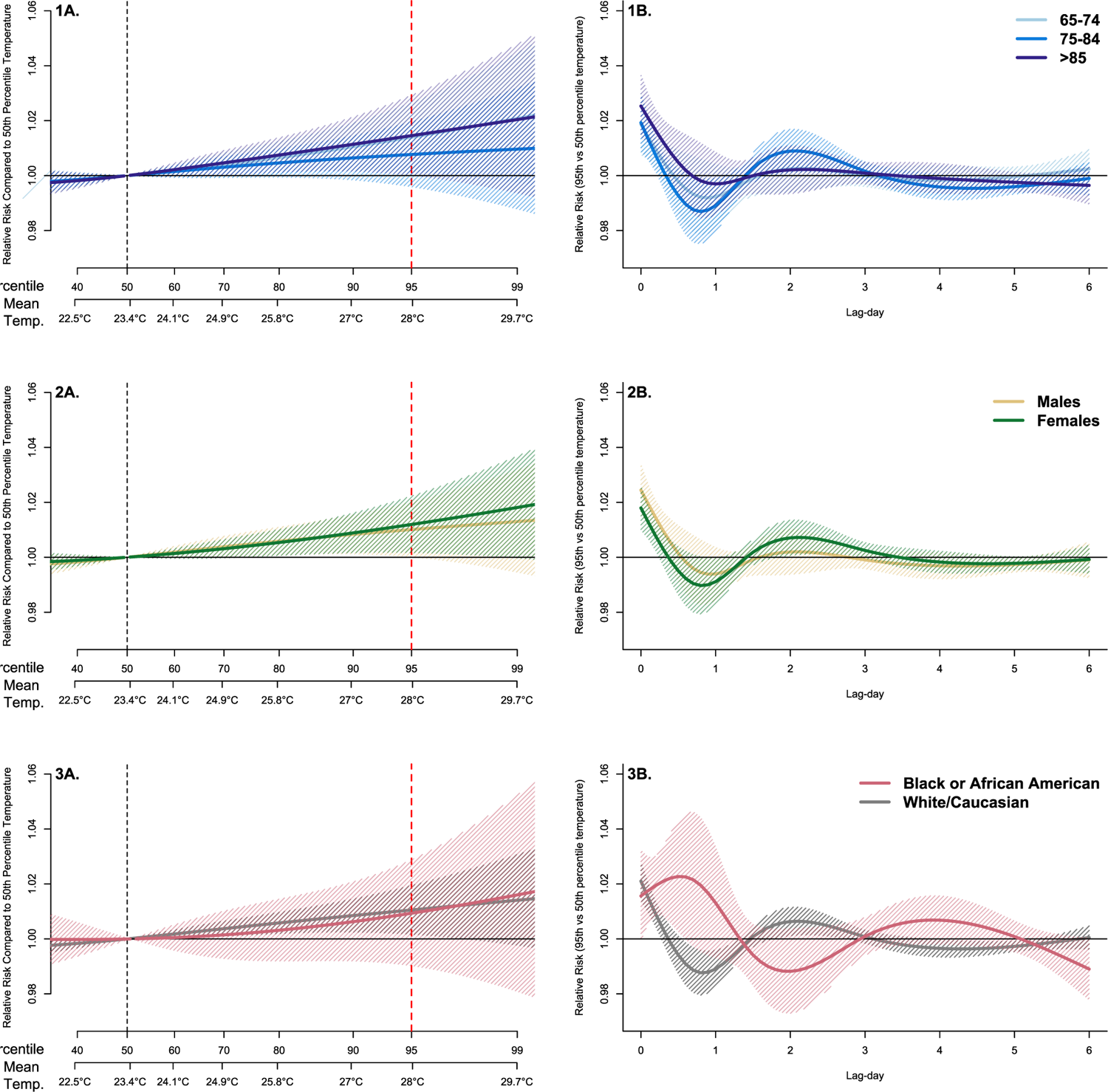
RRs and 95% CIs between ambient mean temperature and all-cause respiratory hospitalizations by age, sex, race/ethnicity, pooled across 120 metropolitan areas. Panels 1A, 2A, and 3A show the overall 7-day cumulative, pooled exposure-response relationship between increases in daily mean temperature and all-cause respiratory hospitalizations by age (1A), sex (2A), and race/ethnicity (3A). Panels 1B, 2B, and 3B demonstrate the pooled lag-response association for each lag day comparing a day of high warm-season temperature (95^th^ percentile) to median temperature (reference exposure). Dashed black line indicates the temperature percentile value used as the centering point for temperature contrasts. The dotted red line indicates the 95th percentile (extreme heat).

**Figure E6.**
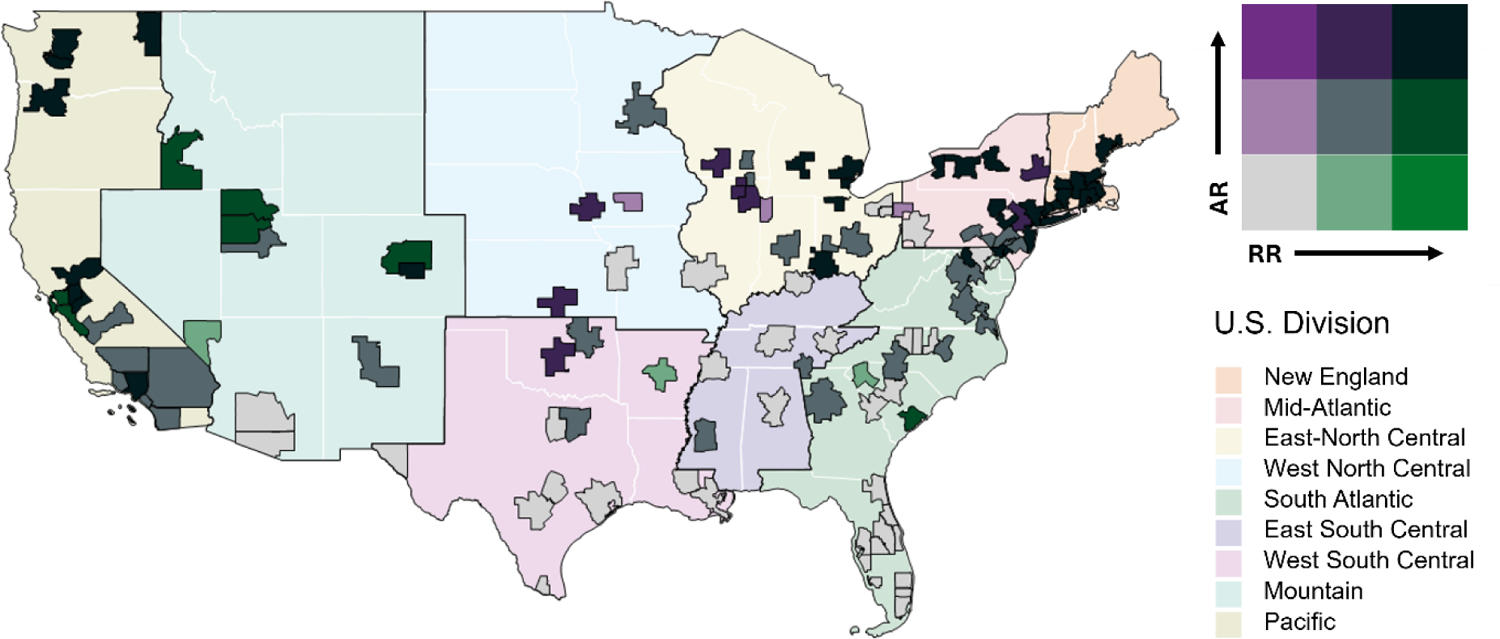
Geographic variation in the risk and burden of 120 metropolitan areas, June-September, 2000-2017. Bi-chrome map (tertile breaks) showing the average relative risk (RR) in each study area by the average annual attributable rate (AR). AR is the annual average warm-season excess number of heat-related all-cause respiratory hospitalizations per 100,000 beneficiaries for each study area.

**Figure E7.**
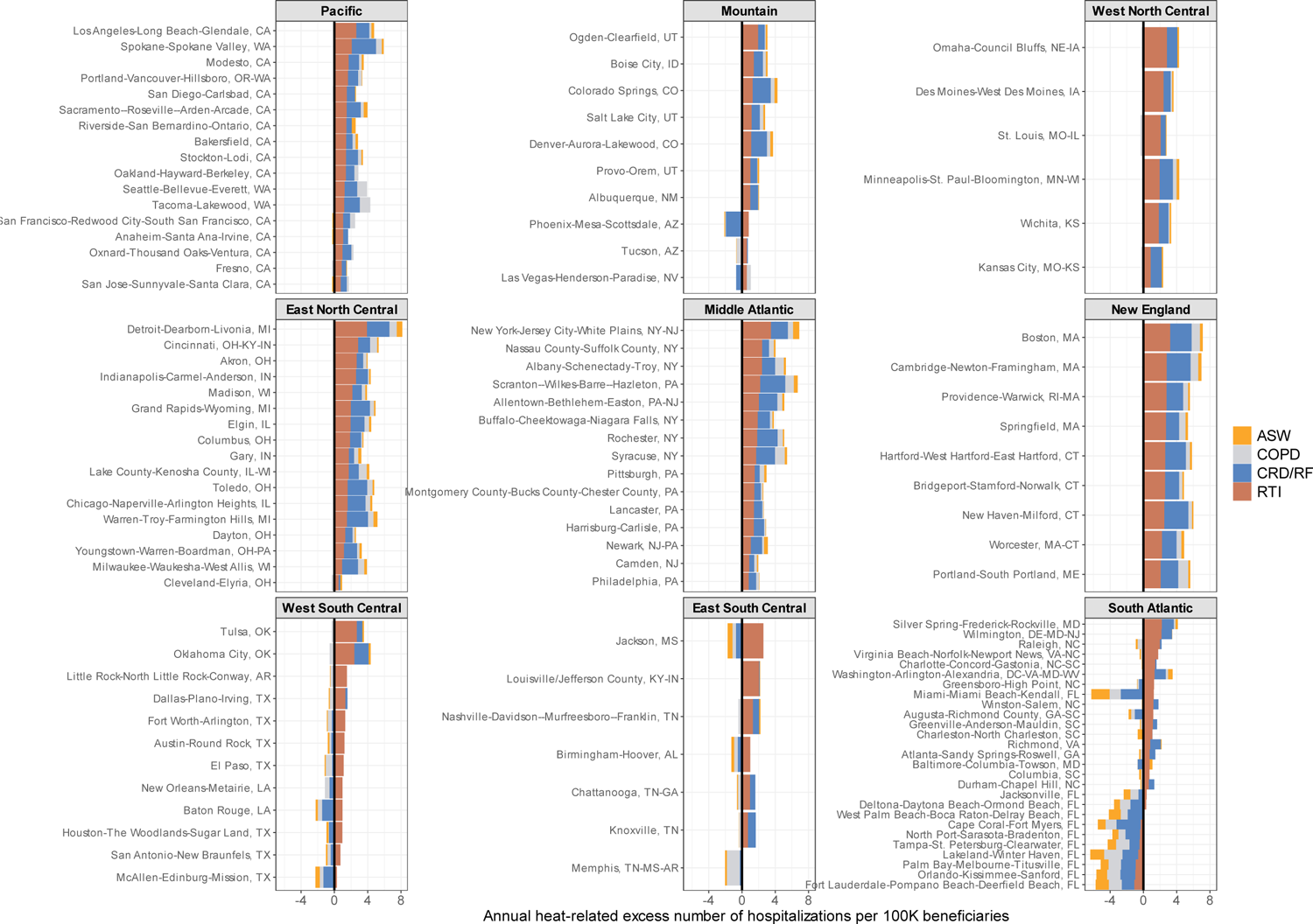
Metropolitan area-specific annual attributable number of excess hospitalizations per 100,000 beneficiaries (annual attributable rate) for each cause-specific outcome by US division.

**Figure E8.**
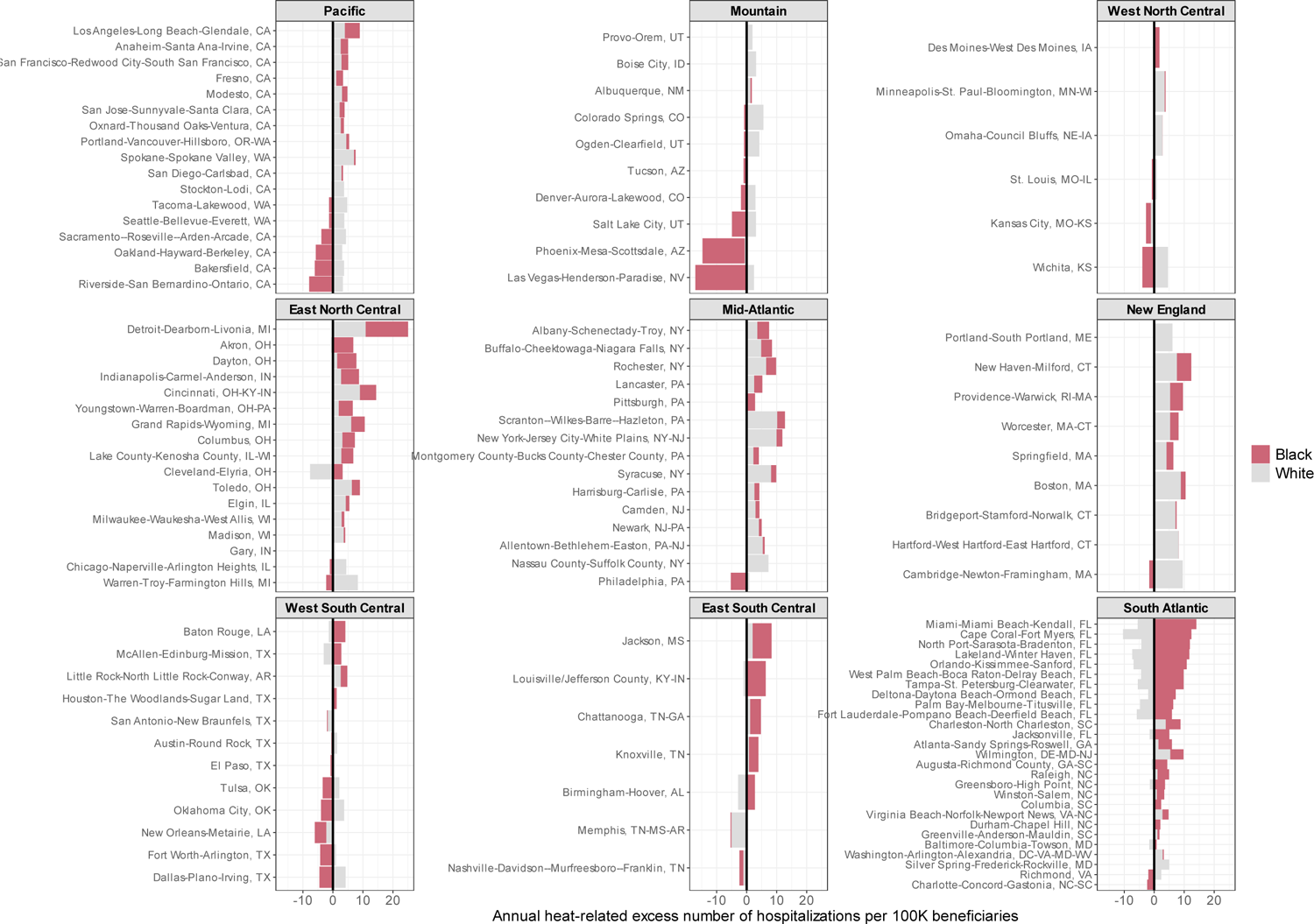
Metropolitan area-specific annual attributable number of excess hospitalizations per 100,000 beneficiaries (annual attributable rate) among black and white beneficiaries for each US division.

**Figure E9.**
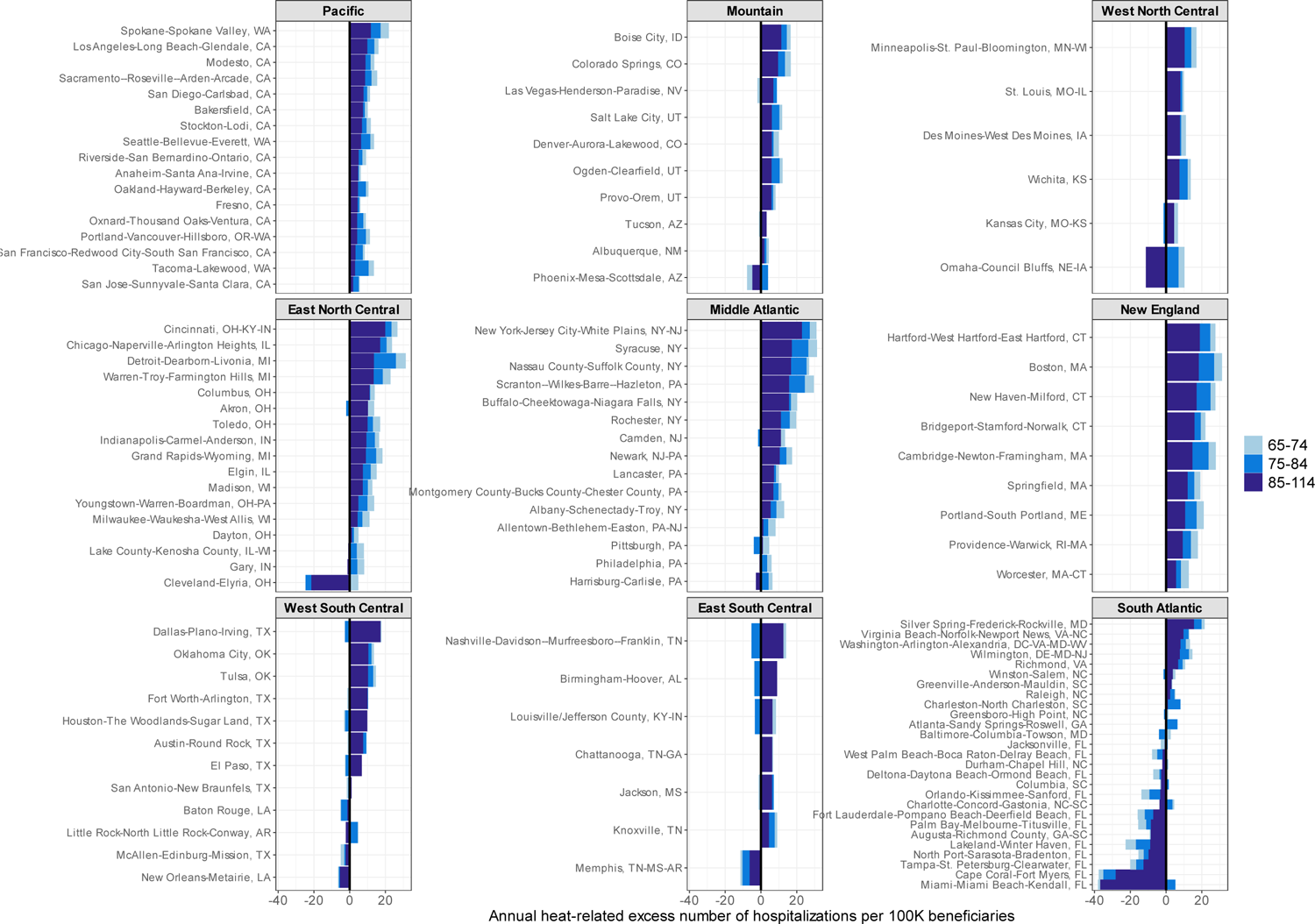
Metropolitan area-specific annual attributable number of excess hospitalizations per 100,000 beneficiaries (annual attributable rate) among beneficiaries across age groups for each US division.

**Figure E10.**
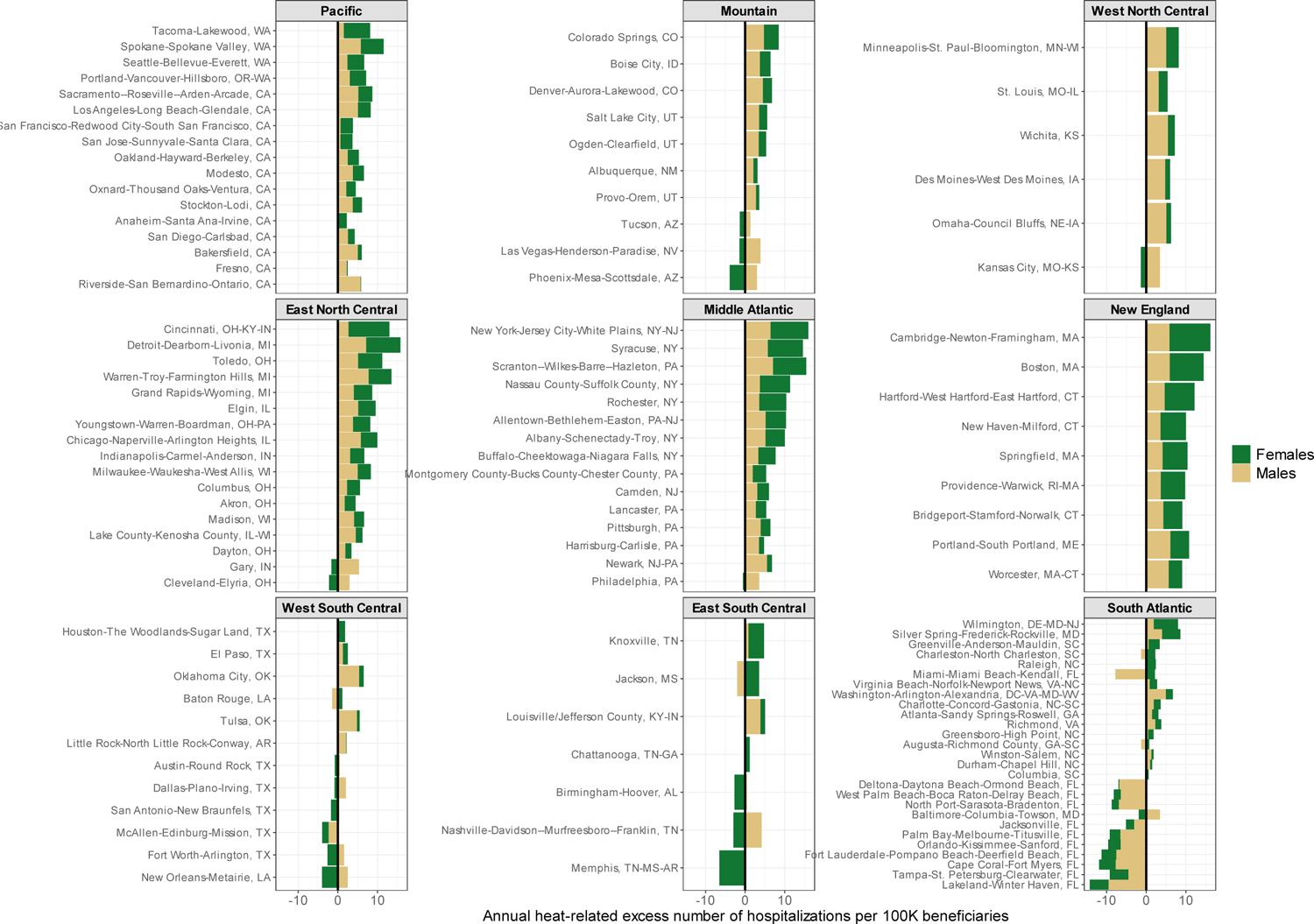
Metropolitan area-specific annual attributable number of excess hospitalizations per 100,000 beneficiaries (annual attributable rate) among female and male beneficiaries for each US division.

**Figure E11.**
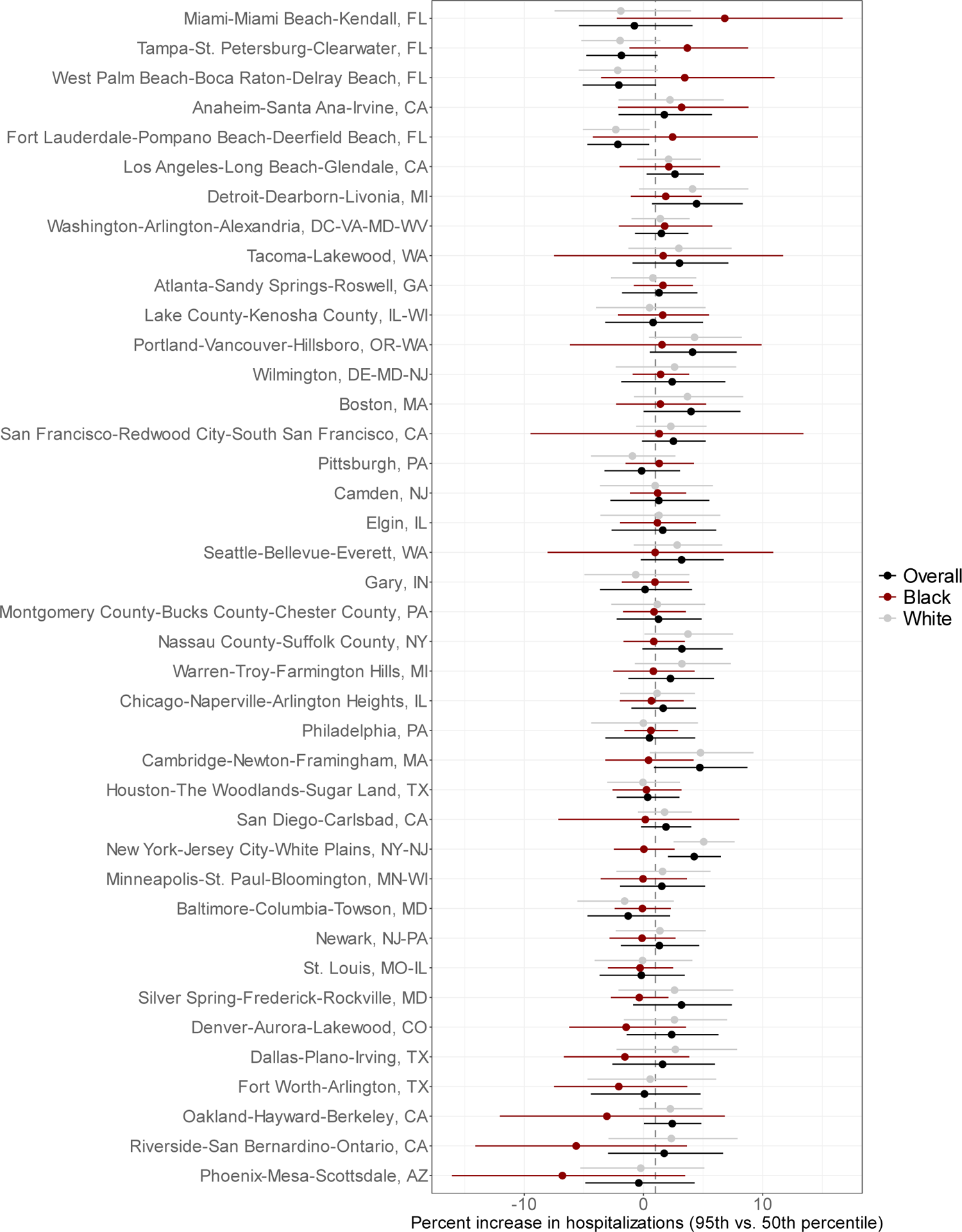
Cumulative percent increase in heat-related all cause respiratory hospitalizations by the overall population, and black and white beneficiaries for the 40 largest metropolitan areas by total population, June-September, 2000-2017. Cumulative associations compare a day of high warm-season temperature (95^th^ percentile) to median temperature (reference exposure). Associations for the overall population are reported in black, associations among black beneficiaries are reported in red, and associations among white beneficiaries are reported in grey.

**Table E1:** International Classification of Disease codes, 9^th^ and 10^th^ revision, for respiratory disease outcomes.

| Outcome | ICD-9 Codes | ICD-10 Codes | Total # primary events, warm-season |
| --- | --- | --- | --- |
| All-cause respiratory diseases | 460-519 | J00-J99 | 3,275,033 |
| Asthma | 493 | J45 | 161,392 |
| Chronic Obstructive Pulmonary Disease | 491, 492, 496 | J41-J44 | 763,628 |
| Respiratory tract infections | 460-466; 480-488 | J00-J18; J20, J21, J22 | 1,276,350 |
| Pneumonia without flu | 480-486 | J12-J18 | 1,203,135 |
| Chronic respiratory (inflammation and fibrotic) diseases/respiratory failure* | 470- 478; 500-519; 490, 494, 495 | J30-J40; J46-J49; J60-J70; J80-J86; J90-J99 | 1,073,663 |

\*Nearly 90% of the events within this category are from respiratory failure, aspiration pneumonitis, pleurisy, pneumothorax, and post-inflammatory pulmonary fibrosis or other interstitial pulmonary diseases (see Table E2)

**Table E2:**
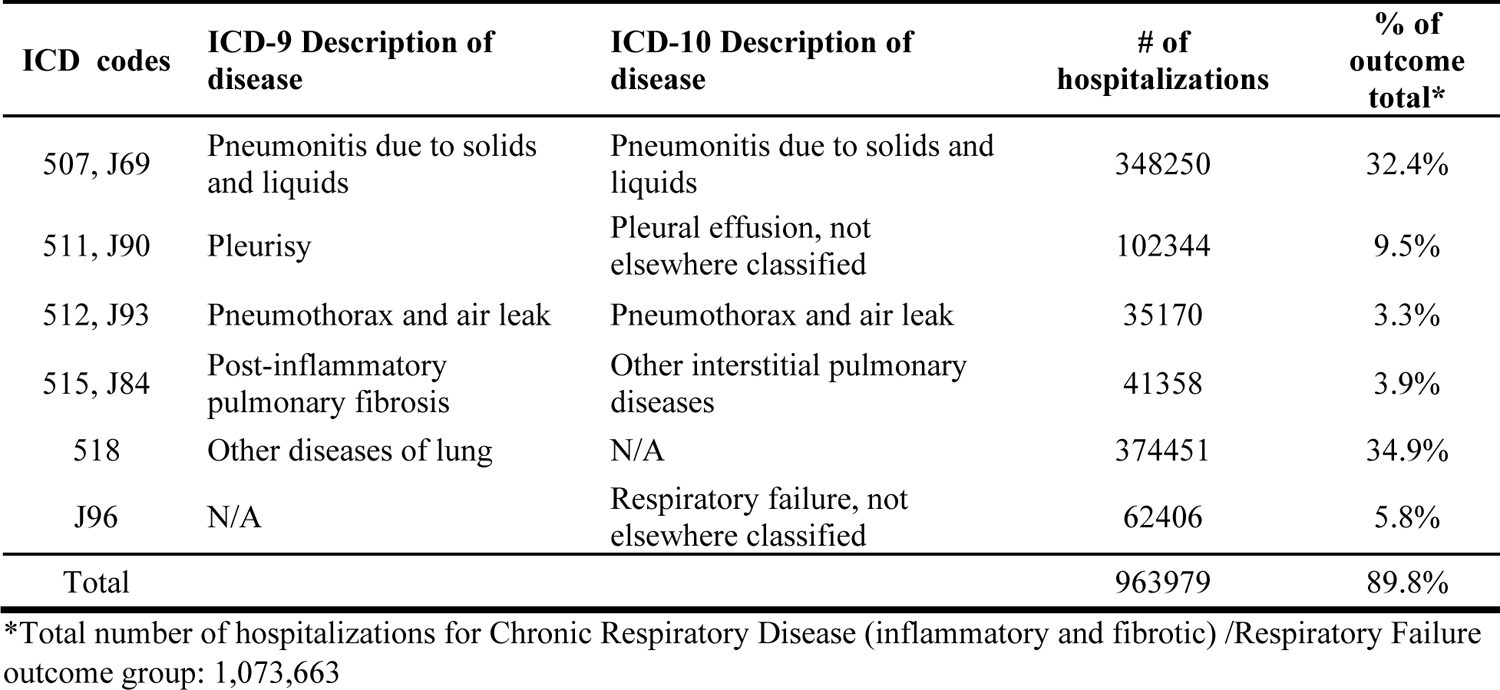
Diagnostic groups with the largest number of events within the Chronic Respiratory Disease (inflammatory and fibrotic) /Respiratory Failure outcome group.

**Table E3.** Descriptive statistics for daily mean temperature averaged across all metropolitan areas, US Divisions, and DOE climate zones, warm season, 2000-2017.

| Geographical Category | Count* | Daily Mean Temperature (°C) |  |  |  |  |  |  |  |  |  |  | Delta^ |
| --- | --- | --- | --- | --- | --- | --- | --- | --- | --- | --- | --- | --- | --- |
|  |  | Min | 25th | Mean | Median | 75th | 95th | 99th | Max | SD | IQR | Range |  |
| All locations | 120 | 0.1 | 20.1 | 23.1 | 23.3 | 26.3 | 29.8 | 33.4 | 42.6 | 4.5 | 6.2 | 42.5 | 6.5 |
| **Division** |  |  |  |  |  |  |  |  |  |  |  |  | 6.5 |
| Pacific | 17 | 2.2 | 18.4 | 21.4 | 21.1 | 24.1 | 29.2 | 33.0 | 42.4 | 4.4 | 5.6 | 40.2 | 8.1 |
| Mountain | 10 | 0.1 | 20.0 | 25.2 | 25.0 | 31.1 | 35.8 | 37.9 | 42.6 | 6.9 | 11.1 | 42.5 | 10.8 |
| West North Central | 6 | 3.9 | 20.1 | 22.7 | 23.0 | 25.8 | 29.3 | 31.4 | 37.0 | 4.4 | 5.7 | 33.1 | 6.3 |
| East North Central | 17 | 5.9 | 18.9 | 21.2 | 21.5 | 23.9 | 26.7 | 28.6 | 33.5 | 3.8 | 5.1 | 27.5 | 5.1 |
| Mid-Atlantic | 15 | 4.6 | 19.1 | 21.4 | 21.7 | 24.0 | 27.0 | 29.1 | 33.6 | 3.7 | 5.0 | 29.0 | 5.2 |
| New England | 9 | 2.5 | 17.8 | 20.2 | 20.5 | 22.9 | 25.9 | 27.8 | 32.1 | 3.7 | 5.1 | 29.5 | 5.4 |
| West South Central | 12 | 6.6 | 25.8 | 27.3 | 27.7 | 29.3 | 31.4 | 33.1 | 37.8 | 2.9 | 3.5 | 31.2 | 3.8 |
| East South Central | 7 | 7.8 | 23.0 | 24.6 | 25.0 | 26.7 | 29.0 | 30.6 | 34.3 | 3.1 | 3.7 | 26.5 | 4.0 |
| South Atlantic | 27 | 7.1 | 23.5 | 25.2 | 25.7 | 27.4 | 29.2 | 30.3 | 37.2 | 3.1 | 4.0 | 30.1 | 3.5 |
| **Climate Zones** |  |  |  |  |  |  |  |  |  |  |  |  |  |
| Cold | 59 | 0.1 | 18.6 | 21.3 | 21.5 | 24.1 | 27.6 | 30.8 | 42.4 | 4.2 | 5.5 | 42.3 | 6.1 |
| Hot-Dry | 13 | 10.2 | 20.5 | 24.4 | 23.3 | 27.8 | 34.5 | 37.1 | 42.6 | 5.2 | 7.3 | 32.4 | 11.2 |
| Hot-Humid | 23 | 6.6 | 26.1 | 27.3 | 27.5 | 28.8 | 30.7 | 32.4 | 37.0 | 2.3 | 2.7 | 30.4 | 3.2 |
| Marine | 1 | 10.6 | 15.9 | 17.5 | 17.2 | 18.7 | 22.0 | 24.9 | 31.8 | 2.4 | 2.9 | 21.2 | 4.8 |
| Mixed-Humid | 24 | 7.8 | 22.3 | 24.2 | 24.5 | 26.5 | 29.3 | 31.4 | 37.8 | 3.4 | 4.2 | 30.0 | 4.8 |

\* The number of metropolitan areas within each grouping
^ Delta is the absolute increase in temperature from the 50<sup>th</sup> percentile to the 95<sup>th</sup> percentile
Abbreviations: 25th: 25th percentile of the temperature distribution; 75th: 75th percentile of the warm-season temperature distribution; 95th: 95th percentile of the warm-season temperature distribution; 99th: 99th percentile of the warm-season temperature distribution; IQR: Interquartile range temperature; Max: Maximum temperature; Min: Minimum temperature; SD: Standard deviation temperature

**Table E4:** Summary of Spearman correlations between daily mean temperature and meteorological variables for each metropolitan area, warm season, 2000-2017.

| Meteorological Variables | n.obs | Spearman correlation with daily mean temperature, summary statistics ($\rho$) | | | | |
| --- | --- | --- | --- | --- | --- | --- |
|  |  | Min | Mean | Median | Max | SD |
| Max Temperature | 120 | 0.62 | 0.90 | 0.92 | 0.95 | 0.07 |
| Min Temperature | 120 | 0.50 | 0.82 | 0.85 | 0.92 | 0.08 |
| Mean Dew Point Temperature | 120 | -0.42 | 0.56 | 0.67 | 0.87 | 0.29 |
| Mean Humidity* | 120 | -0.71 | -0.28 | -0.24 | 0.10 | 0.25 |
| Max Heat Index | 120 | 0.33 | 0.85 | 0.91 | 0.94 | 0.14 |
| Mean Heat Index | 120 | 0.89 | 0.99 | 1.00 | 1.00 | 0.02 |

\*Humidity values were adjusted so that no values were below zero percent or above 100%
Abbreviations: Max: Maximum temperature; Min: Minimum temperature; n.obs: Number of metropolitan areas contributing Spearman Correlation data; SD: Standard deviation temperature

**Table E5.** Minimum hospitalization percentiles, Minimum hospitalization temperatures, and I^2^ heterogeneity values from meta-regression models, overall and by sub-group.

| Outcome | MHP | MHT | $I^2$ Statistic |
| --- | --- | --- | --- |
| **All-cause respiratory** |  |  |  |
| Mean Temp, lag 0-6 (primary) | 22% | 20.7 | 8.7% |
| Mean Temp., lag 0-3 (sensitivity) | 2% | 15.8 | 11.3% |
| Min. Temp. (sensitivity) | 36% | 16.4 | 2.4% |
| Max. Temp. (sensitivity) | 20% | 26.6 | 12.3% |
| Mean HI (sensitivity) | 17% | 20.0 | 3.4% |
| Max. HI (sensitivity) | 5% | 24.2 | 15.9% |
| **Age Group** |  |  |  |
| 65-74 | 11% | 19.0 | 0.0% |
| 75-84 | 19% | 20.3 | 3.9% |
| 85-114 | 25% | 21.1 | 0.6% |
| **Sex** |  |  |  |
| Males | 22% | 20.7 | 3.8% |
| Females | 21% | 20.6 | 0.0% |
| **Race/Ethnicity** |  |  |  |
| Black | 39% | 22.5 | 1.4% |
| White | 20% | 20.5 | 10.5% |
| **Cause-Specific Outcomes** |  |  |  |
| Asthma | 0% | 10.1 | 0.0% |
| COPD | 0% | 10.1 | 12.4% |
| CRD/RF | 28% | 21.4 | 5.9% |
| RTI | 20% | 20.5 | 4.8% |

**Table E6.** Metropolitan area daily ambient temperature summary statistics, warm season (June-September), 2000-2017.

| Study Location | Division | Daily Mean Temperature (°C) |  |  |  |  |  |  |  |  |  |  |
| --- | --- | --- | --- | --- | --- | --- | --- | --- | --- | --- | --- | --- |
|  |  | Min | 25th | Mean | Median | 95th | 99th | Max | SD | IQR | Range | Delta* |
| Akron, OH | East North Central | 8.04 | 18.50 | 20.83 | 21.17 | 26.15 | 27.61 | 30.00 | 3.60 | 5.02 | 21.96 | 4.98 |
| Albany-Schenectady-Troy, NY | Mid-Atlantic | 4.68 | 17.18 | 19.51 | 19.82 | 24.96 | 26.74 | 31.30 | 3.67 | 4.94 | 26.62 | 5.13 |
| Albuquerque, NM | Mountain | 6.93 | 20.14 | 22.29 | 22.60 | 27.29 | 28.80 | 32.28 | 3.35 | 4.53 | 25.35 | 4.69 |
| Allentown-Bethlehem-Easton, PA-NJ | Mid-Atlantic | 7.11 | 19.12 | 21.32 | 21.64 | 26.62 | 28.48 | 32.44 | 3.55 | 4.74 | 25.33 | 4.99 |
| Anaheim-Santa Ana-Irvine, CA | Pacific | 14.63 | 19.98 | 21.81 | 21.63 | 26.22 | 28.30 | 31.25 | 2.51 | 3.45 | 16.61 | 4.59 |
| Atlanta-Sandy Springs-Roswell, GA | South Atlantic | 12.55 | 23.56 | 24.93 | 25.14 | 28.71 | 29.90 | 33.09 | 2.58 | 3.07 | 20.54 | 3.57 |
| Augusta-Richmond County, GA-SC | South Atlantic | 14.46 | 24.38 | 25.74 | 25.86 | 29.65 | 30.89 | 32.83 | 2.52 | 2.98 | 18.37 | 3.79 |
| Austin-Round Rock, TX | West South Central | 14.14 | 26.27 | 27.76 | 28.07 | 31.25 | 32.40 | 35.82 | 2.51 | 3.29 | 21.69 | 3.18 |
| Bakersfield, CA | Pacific | 10.74 | 23.64 | 26.30 | 26.41 | 32.67 | 34.94 | 40.24 | 3.98 | 5.42 | 29.51 | 6.25 |
| Baltimore-Columbia-Towson, MD | South Atlantic | 8.60 | 21.50 | 23.55 | 23.82 | 28.65 | 30.61 | 33.54 | 3.38 | 4.38 | 24.94 | 4.83 |
| Baton Rouge, LA | West South Central | 14.83 | 25.70 | 26.77 | 26.87 | 29.63 | 30.75 | 32.27 | 1.95 | 2.38 | 17.43 | 2.76 |
| Birmingham-Hoover, AL | East South Central | 12.57 | 24.01 | 25.27 | 25.57 | 28.88 | 30.20 | 31.75 | 2.53 | 2.88 | 19.18 | 3.31 |
| Boise City, ID | Mountain | 6.65 | 18.20 | 21.39 | 21.95 | 28.16 | 30.27 | 34.56 | 4.71 | 6.77 | 27.92 | 6.21 |
| Boston, MA | New England | 6.23 | 17.76 | 20.20 | 20.43 | 25.96 | 27.73 | 31.62 | 3.72 | 5.18 | 25.39 | 5.53 |
| Bridgeport-Stamford-Norwalk, CT | New England | 6.97 | 19.17 | 21.30 | 21.64 | 26.34 | 28.41 | 31.50 | 3.41 | 4.50 | 24.54 | 4.70 |
| Buffalo-Cheektowaga-Niagara Falls, NY | Mid-Atlantic | 6.24 | 17.74 | 20.05 | 20.27 | 25.56 | 27.29 | 29.90 | 3.60 | 4.95 | 23.66 | 5.29 |
| Cambridge-Newton-Framingham, MA | New England | 5.43 | 17.61 | 20.14 | 20.41 | 26.10 | 28.04 | 32.06 | 3.84 | 5.32 | 26.63 | 5.69 |
| Camden, NJ | Mid-Atlantic | 10.89 | 20.85 | 22.96 | 23.25 | 28.32 | 30.31 | 32.97 | 3.46 | 4.44 | 22.08 | 5.07 |
| Cape Coral-Fort Myers, FL | South Atlantic | 22.37 | 26.58 | 27.38 | 27.44 | 29.23 | 29.75 | 30.49 | 1.19 | 1.65 | 8.12 | 1.79 |
| Charleston-North Charleston, SC | South Atlantic | 15.79 | 24.99 | 26.16 | 26.33 | 29.45 | 30.62 | 31.81 | 2.18 | 2.58 | 16.02 | 3.12 |
| Charlotte-Concord-Gastonia, NC-SC | South Atlantic | 11.03 | 22.74 | 24.21 | 24.49 | 28.36 | 29.88 | 33.47 | 2.83 | 3.33 | 22.44 | 3.87 |
| Chattanooga, TN-GA | East South Central | 11.88 | 22.82 | 24.24 | 24.59 | 27.98 | 29.19 | 31.66 | 2.68 | 3.21 | 19.78 | 3.39 |
| Chicago-Naperville-Arlington Heights, IL | East North Central | 7.68 | 19.03 | 21.42 | 21.78 | 27.04 | 29.24 | 32.80 | 3.84 | 5.15 | 25.11 | 5.26 |
| Cincinnati, OH-KY-IN | East North Central | 8.69 | 20.33 | 22.39 | 22.82 | 27.13 | 28.85 | 32.33 | 3.39 | 4.49 | 23.64 | 4.31 |
| Cleveland-Elyria, OH | East North Central | 7.76 | 18.75 | 21.02 | 21.32 | 26.40 | 28.00 | 30.76 | 3.58 | 4.87 | 23.00 | 5.08 |
| Colorado Springs, CO | Mountain | 0.49 | 16.90 | 19.25 | 19.65 | 24.98 | 26.64 | 30.79 | 3.91 | 5.12 | 30.30 | 5.33 |
| Columbia, SC | South Atlantic | 13.82 | 24.39 | 25.79 | 25.96 | 29.82 | 31.21 | 33.77 | 2.61 | 3.07 | 19.95 | 3.85 |
| Columbus, OH | East North Central | 7.85 | 19.35 | 21.53 | 21.95 | 26.33 | 27.94 | 31.33 | 3.45 | 4.76 | 23.48 | 4.38 |
| Dallas-Plano-Irving, TX | West South Central | 13.04 | 25.79 | 27.79 | 28.03 | 32.40 | 33.75 | 36.47 | 3.12 | 4.23 | 23.43 | 4.37 |
| Dayton, OH | East North Central | 8.17 | 19.75 | 21.87 | 22.31 | 26.76 | 28.58 | 32.07 | 3.51 | 4.72 | 23.89 | 4.45 |
| Deltona-Daytona Beach-Ormond Beach, FL | South Atlantic | 21.07 | 26.13 | 27.01 | 27.02 | 29.24 | 29.92 | 31.50 | 1.37 | 1.80 | 10.43 | 2.22 |
| Denver-Aurora-Lakewood, CO | Mountain | 0.10 | 15.94 | 18.53 | 18.88 | 24.77 | 26.51 | 32.49 | 4.20 | 5.61 | 32.39 | 5.90 |
| Des Moines-West Des Moines, IA | West North Central | 6.31 | 19.46 | 21.83 | 22.16 | 27.85 | 29.65 | 32.20 | 4.00 | 5.19 | 25.89 | 5.69 |
| Detroit-Dearborn-Livonia, MI | East North Central | 6.95 | 18.75 | 21.08 | 21.37 | 26.62 | 28.69 | 32.06 | 3.75 | 5.05 | 25.11 | 5.25 |
| Durham-Chapel Hill, NC | South Atlantic | 11.86 | 22.64 | 24.28 | 24.61 | 28.57 | 30.01 | 32.49 | 2.96 | 3.67 | 20.63 | 3.97 |
| Elgin, IL | East North Central | 7.26 | 18.61 | 20.99 | 21.32 | 26.65 | 28.67 | 32.25 | 3.84 | 5.18 | 24.99 | 5.33 |
| El Paso, TX | West South Central | 12.51 | 24.67 | 26.70 | 26.91 | 31.57 | 33.00 | 35.01 | 3.19 | 4.34 | 22.50 | 4.66 |
| Fort Lauderdale-Pompano Beach-Deerfield Beach, FL | South Atlantic | 22.94 | 27.31 | 28.11 | 28.23 | 29.92 | 30.34 | 33.33 | 1.24 | 1.73 | 10.39 | 1.69 |
| Fort Worth-Arlington, TX | West South Central | 13.27 | 26.03 | 28.07 | 28.36 | 32.79 | 34.15 | 36.98 | 3.22 | 4.37 | 23.72 | 4.43 |
| Fresno, CA | Pacific | 13.45 | 23.59 | 25.90 | 26.12 | 31.39 | 33.17 | 37.55 | 3.45 | 4.70 | 24.10 | 5.27 |
| Gary, IN | East North Central | 8.57 | 19.33 | 21.64 | 22.02 | 27.06 | 29.04 | 32.66 | 3.74 | 5.00 | 24.08 | 5.05 |
| Grand Rapids-Wyoming, MI | East North Central | 5.94 | 17.46 | 19.92 | 20.25 | 25.66 | 27.94 | 31.04 | 3.84 | 5.20 | 25.10 | 5.41 |
| Greensboro-High Point, NC | South Atlantic | 11.09 | 22.26 | 23.88 | 24.24 | 28.12 | 29.65 | 32.64 | 2.96 | 3.62 | 21.54 | 3.88 |
| Greenville-Anderson-Mauldin, SC | South Atlantic | 12.15 | 23.27 | 24.71 | 24.93 | 28.82 | 30.26 | 33.21 | 2.74 | 3.21 | 21.06 | 3.88 |
| Harrisburg-Carlisle, PA | Mid-Atlantic | 8.48 | 19.58 | 21.74 | 22.07 | 26.93 | 28.75 | 32.23 | 3.50 | 4.64 | 23.75 | 4.86 |
| Hartford-West Hartford-East Hartford, CT | New England | 6.26 | 18.51 | 20.81 | 21.16 | 26.23 | 28.13 | 31.85 | 3.58 | 4.84 | 25.59 | 5.07 |
| Houston-The Woodlands-Sugar Land, TX | West South Central | 14.44 | 26.60 | 27.77 | 28.10 | 30.44 | 31.35 | 34.26 | 1.98 | 2.54 | 19.82 | 2.34 |
| Indianapolis-Carmel-Anderson, IN | East North Central | 8.51 | 20.12 | 22.27 | 22.70 | 27.28 | 29.10 | 33.45 | 3.55 | 4.66 | 24.95 | 4.58 |
| Jackson, MS | East South Central | 14.72 | 24.82 | 26.09 | 26.29 | 29.60 | 30.57 | 32.23 | 2.38 | 2.87 | 17.51 | 3.31 |
| Jacksonville, FL | South Atlantic | 17.77 | 25.78 | 26.72 | 26.73 | 29.23 | 30.17 | 33.21 | 1.55 | 1.94 | 15.44 | 2.49 |
| Kansas City, MO-KS | West North Central | 8.87 | 21.50 | 24.05 | 24.33 | 30.20 | 32.19 | 35.16 | 4.05 | 5.40 | 26.28 | 5.87 |
| Knoxville, TN | East South Central | 9.34 | 21.62 | 23.13 | 23.50 | 26.91 | 28.36 | 30.84 | 2.75 | 3.35 | 21.49 | 3.40 |
| Lake County-Kenosha County, IL-WI | East North Central | 8.11 | 17.97 | 20.52 | 20.80 | 26.34 | 29.09 | 32.37 | 3.90 | 5.34 | 24.27 | 5.54 |
| Lakeland-Winter Haven, FL | South Atlantic | 22.17 | 26.34 | 27.22 | 27.30 | 29.31 | 30.04 | 31.74 | 1.33 | 1.81 | 9.56 | 2.00 |
| Lancaster, PA | Mid-Atlantic | 9.54 | 20.27 | 22.40 | 22.70 | 27.61 | 29.39 | 33.07 | 3.47 | 4.56 | 23.53 | 4.90 |
| Las Vegas-Henderson-Paradise, NV | Mountain | 12.86 | 28.48 | 30.98 | 31.51 | 36.55 | 38.41 | 42.37 | 3.89 | 5.34 | 29.51 | 5.05 |
| Little Rock-North Little Rock-Conway, AR | West South Central | 12.22 | 24.07 | 26.00 | 26.24 | 30.57 | 32.20 | 34.76 | 3.14 | 4.15 | 22.54 | 4.34 |
| Los Angeles-Long Beach-Glendale, CA | Pacific | 12.53 | 20.23 | 22.24 | 21.98 | 27.32 | 29.69 | 35.50 | 2.84 | 3.82 | 22.97 | 5.34 |
| Louisville/Jefferson County, KY-IN | East South Central | 9.56 | 21.65 | 23.57 | 23.98 | 28.29 | 30.13 | 33.12 | 3.39 | 4.27 | 23.55 | 4.31 |
| Madison, WI | East North Central | 6.33 | 17.94 | 20.35 | 20.65 | 26.17 | 28.65 | 32.64 | 3.99 | 5.23 | 26.31 | 5.51 |
| McAllen-Edinburg-Mission, TX | West South Central | 18.05 | 28.09 | 29.04 | 29.26 | 31.56 | 32.35 | 34.43 | 1.83 | 2.23 | 16.38 | 2.30 |
| Memphis, TN-MS-AR | East South Central | 13.24 | 23.92 | 25.71 | 26.02 | 30.19 | 31.58 | 34.31 | 3.08 | 3.94 | 21.07 | 4.17 |
| Miami-Miami Beach-Kendall, FL | South Atlantic | 23.10 | 27.26 | 28.01 | 28.12 | 29.73 | 30.19 | 31.31 | 1.16 | 1.61 | 8.21 | 1.60 |
| Milwaukee-Waukesha-West Allis, WI | East North Central | 7.33 | 17.60 | 20.11 | 20.37 | 26.02 | 28.84 | 32.38 | 3.93 | 5.26 | 25.05 | 5.65 |
| Minneapolis-St. Paul-Bloomington, MN-WI | West North Central | 3.94 | 17.86 | 20.25 | 20.65 | 26.29 | 28.77 | 32.48 | 4.17 | 5.31 | 28.53 | 5.64 |
| Modesto, CA | Pacific | 10.16 | 21.24 | 23.45 | 23.29 | 29.34 | 31.67 | 35.92 | 3.33 | 4.27 | 25.77 | 6.05 |
| Montgomery County-Bucks County-Chester County, PA | Mid-Atlantic | 9.20 | 20.29 | 22.44 | 22.72 | 27.80 | 29.76 | 33.42 | 3.51 | 4.58 | 24.21 | 5.07 |
| Nashville-Davidson--Murfreesboro--Franklin, TN | East South Central | 7.77 | 22.94 | 24.67 | 25.06 | 29.10 | 30.71 | 33.67 | 3.14 | 3.86 | 25.90 | 4.04 |
| Nassau County-Suffolk County, NY | Mid-Atlantic | 9.34 | 19.85 | 21.95 | 22.24 | 27.12 | 29.16 | 32.49 | 3.34 | 4.35 | 23.14 | 4.87 |
| Newark, NJ-PA | Mid-Atlantic | 6.19 | 19.54 | 21.81 | 22.11 | 27.29 | 29.32 | 33.57 | 3.59 | 4.75 | 27.38 | 5.18 |
| New Haven-Milford, CT | New England | 6.96 | 18.78 | 20.91 | 21.28 | 25.95 | 27.90 | 30.96 | 3.42 | 4.56 | 24.00 | 4.67 |
| New Orleans-Metairie, LA | West South Central | 6.63 | 26.49 | 27.43 | 27.56 | 30.00 | 30.92 | 32.36 | 1.73 | 2.07 | 25.73 | 2.44 |
| New York-Jersey City-White Plains, NY-NJ | Mid-Atlantic | 6.36 | 19.89 | 22.10 | 22.40 | 27.54 | 29.60 | 33.57 | 3.53 | 4.60 | 27.21 | 5.14 |
| North Port-Sarasota-Bradenton, FL | South Atlantic | 22.57 | 26.63 | 27.41 | 27.48 | 29.29 | 29.97 | 31.22 | 1.22 | 1.64 | 8.65 | 1.81 |
| Oakland-Hayward-Berkeley, CA | Pacific | 12.30 | 17.37 | 19.24 | 18.89 | 24.26 | 26.98 | 34.97 | 2.66 | 3.33 | 22.67 | 5.38 |
| Ogden-Clearfield, UT | Mountain | 3.41 | 18.63 | 21.50 | 22.28 | 27.93 | 29.51 | 32.07 | 4.69 | 6.33 | 28.66 | 5.65 |
| Oklahoma City, OK | West South Central | 9.94 | 23.90 | 26.23 | 26.47 | 31.80 | 33.63 | 37.79 | 3.71 | 4.97 | 27.85 | 5.32 |
| Omaha-Council Bluffs, NE-IA | West North Central | 7.03 | 19.96 | 22.45 | 22.81 | 28.59 | 30.37 | 32.92 | 4.16 | 5.48 | 25.89 | 5.78 |
| Orlando-Kissimmee-Sanford, FL | South Atlantic | 19.95 | 26.22 | 27.08 | 27.13 | 29.21 | 29.92 | 31.84 | 1.32 | 1.76 | 11.89 | 2.08 |
| Oxnard-Thousand Oaks-Ventura, CA | Pacific | 13.34 | 18.60 | 20.29 | 20.10 | 24.62 | 26.91 | 32.30 | 2.43 | 3.16 | 18.96 | 4.52 |
| Palm Bay-Melbourne-Titusville, FL | South Atlantic | 22.09 | 26.42 | 27.23 | 27.30 | 29.14 | 29.68 | 31.57 | 1.24 | 1.68 | 9.48 | 1.84 |
| Philadelphia, PA | Mid-Atlantic | 10.80 | 20.87 | 22.96 | 23.28 | 28.32 | 30.19 | 33.26 | 3.47 | 4.44 | 22.46 | 5.04 |
| Phoenix-Mesa-Scottsdale, AZ | Mountain | 19.72 | 30.67 | 32.73 | 33.00 | 37.40 | 38.93 | 42.57 | 3.14 | 4.37 | 22.85 | 4.40 |
| Pittsburgh, PA | Mid-Atlantic | 7.41 | 18.60 | 20.76 | 21.15 | 25.73 | 27.16 | 31.11 | 3.46 | 4.73 | 23.70 | 4.58 |
| Portland-South Portland, ME | New England | 2.54 | 16.10 | 18.42 | 18.71 | 24.17 | 26.10 | 31.72 | 3.67 | 4.85 | 29.17 | 5.46 |
| Portland-Vancouver-Hillsboro, OR-WA | Pacific | 9.72 | 16.36 | 18.66 | 18.47 | 24.57 | 27.35 | 32.56 | 3.38 | 4.43 | 22.84 | 6.11 |
| Providence-Warwick, RI-MA | New England | 6.64 | 18.25 | 20.45 | 20.69 | 25.67 | 27.54 | 30.87 | 3.48 | 4.69 | 24.23 | 4.97 |
| Provo-Orem, UT | Mountain | 5.28 | 19.45 | 22.12 | 22.90 | 28.21 | 29.77 | 33.60 | 4.42 | 5.91 | 28.32 | 5.32 |
| Raleigh, NC | South Atlantic | 12.84 | 23.04 | 24.69 | 24.99 | 28.99 | 30.37 | 32.25 | 2.91 | 3.63 | 19.41 | 4.00 |
| Richmond, VA | South Atlantic | 10.42 | 22.03 | 23.98 | 24.28 | 28.71 | 30.39 | 34.22 | 3.21 | 4.17 | 23.80 | 4.44 |
| Riverside-San Bernardino-Ontario, CA | Pacific | 12.56 | 23.63 | 26.61 | 26.31 | 34.02 | 36.87 | 42.38 | 4.23 | 5.81 | 29.82 | 7.71 |
| Rochester, NY | Mid-Atlantic | 5.61 | 17.50 | 19.89 | 20.09 | 25.63 | 27.45 | 30.82 | 3.70 | 5.02 | 25.20 | 5.54 |
| Sacramento--Roseville--Arden-Arcade, CA | Pacific | 2.20 | 19.80 | 21.94 | 21.92 | 27.78 | 30.19 | 34.93 | 3.51 | 4.36 | 32.73 | 5.85 |
| St. Louis, MO-IL | West North Central | 9.12 | 21.45 | 23.76 | 24.09 | 29.34 | 31.23 | 34.12 | 3.78 | 4.99 | 24.99 | 5.25 |
| Salt Lake City, UT | Mountain | 5.32 | 19.83 | 22.66 | 23.47 | 29.06 | 30.60 | 34.15 | 4.68 | 6.29 | 28.83 | 5.60 |
| San Antonio-New Braunfels, TX | West South Central | 13.94 | 26.64 | 28.00 | 28.32 | 31.13 | 32.01 | 34.94 | 2.32 | 3.08 | 21.00 | 2.81 |
| San Diego-Carlsbad, CA | Pacific | 13.98 | 20.13 | 22.04 | 21.83 | 26.78 | 29.09 | 37.13 | 2.65 | 3.56 | 23.15 | 4.95 |
| San Francisco-Redwood City-South San Francisco, CA | Pacific | 10.60 | 15.85 | 17.50 | 17.24 | 22.03 | 24.93 | 31.79 | 2.40 | 2.90 | 21.19 | 4.79 |
| San Jose-Sunnyvale-Santa Clara, CA | Pacific | 12.40 | 18.03 | 19.62 | 19.32 | 24.02 | 26.69 | 32.56 | 2.35 | 2.85 | 20.16 | 4.70 |
| Scranton--Wilkes-Barre--Hazleton, PA | Mid-Atlantic | 5.51 | 17.76 | 20.04 | 20.37 | 25.39 | 27.10 | 32.82 | 3.59 | 4.88 | 27.31 | 5.02 |
| Seattle-Bellevue-Everett, WA | Pacific | 7.59 | 14.81 | 16.87 | 16.72 | 22.11 | 24.68 | 29.96 | 2.99 | 3.92 | 22.37 | 5.39 |
| Silver Spring-Frederick-Rockville, MD | South Atlantic | 7.08 | 20.93 | 23.03 | 23.32 | 28.10 | 30.00 | 33.09 | 3.40 | 4.49 | 26.01 | 4.79 |
| Spokane-Spokane Valley, WA | Pacific | 4.29 | 15.36 | 18.68 | 18.90 | 25.69 | 27.52 | 32.33 | 4.46 | 6.62 | 28.04 | 6.79 |
| Springfield, MA | New England | 5.86 | 17.60 | 20.04 | 20.37 | 25.65 | 27.63 | 31.57 | 3.74 | 5.13 | 25.71 | 5.28 |
| Stockton-Lodi, CA | Pacific | 11.29 | 20.29 | 22.46 | 22.18 | 28.18 | 30.92 | 35.68 | 3.22 | 4.17 | 24.39 | 6.00 |
| Syracuse, NY | Mid-Atlantic | 4.60 | 17.16 | 19.57 | 19.80 | 25.30 | 27.12 | 30.44 | 3.73 | 5.04 | 25.84 | 5.51 |
| Tacoma-Lakewood, WA | Pacific | 7.66 | 14.79 | 16.93 | 16.77 | 22.36 | 24.79 | 29.45 | 3.09 | 4.09 | 21.79 | 5.59 |
| Tampa-St. Petersburg-Clearwater, FL | South Atlantic | 19.91 | 26.61 | 27.44 | 27.51 | 29.47 | 30.08 | 32.28 | 1.31 | 1.76 | 12.37 | 1.96 |
| Toledo, OH | East North Central | 7.44 | 18.78 | 21.05 | 21.37 | 26.36 | 28.13 | 32.01 | 3.65 | 4.94 | 24.57 | 4.99 |
| Tucson, AZ | Mountain | 17.53 | 27.27 | 29.28 | 29.36 | 33.88 | 35.53 | 39.91 | 2.95 | 4.14 | 22.38 | 4.52 |
| Tulsa, OK | West South Central | 10.45 | 23.44 | 25.80 | 26.09 | 31.36 | 33.49 | 37.80 | 3.81 | 5.10 | 27.34 | 5.27 |
| Virginia Beach-Norfolk-Newport News, VA-NC | South Atlantic | 12.90 | 23.04 | 24.77 | 24.94 | 29.07 | 30.59 | 33.82 | 2.86 | 3.69 | 20.93 | 4.13 |
| Warren-Troy-Farmington Hills,<br>MI | East North Central | 6.46 | 18.04 | 20.50 | 20.77 | 26.30 | 28.38 | 32.04 | 3.84 | 5.22 | 25.58 | 5.52 |
| Washington-Arlington-Alexandria, DC-VA-MD-WV | South Atlantic | 9.52 | 21.38 | 23.35 | 23.61 | 28.25 | 30.12 | 37.19 | 3.31 | 4.28 | 27.66 | 4.63 |
| West Palm Beach-Boca Raton-Delray Beach, FL | South Atlantic | 22.87 | 27.15 | 28.01 | 28.11 | 30.06 | 30.62 | 33.23 | 1.32 | 1.80 | 10.35 | 1.95 |
| Wichita, KS | West North Central | 7.77 | 22.14 | 24.88 | 25.11 | 31.16 | 32.88 | 37.02 | 4.11 | 5.63 | 29.25 | 6.05 |
| Wilmington, DE-MD-NJ | South Atlantic | 10.80 | 21.16 | 23.23 | 23.52 | 28.40 | 30.31 | 33.22 | 3.39 | 4.37 | 22.42 | 4.88 |
| Winston-Salem, NC | South Atlantic | 10.40 | 21.68 | 23.28 | 23.65 | 27.45 | 29.04 | 32.43 | 2.95 | 3.57 | 22.03 | 3.80 |
| Worcester, MA-CT | New England | 5.40 | 17.69 | 20.14 | 20.46 | 25.78 | 27.80 | 31.35 | 3.75 | 5.18 | 25.95 | 5.32 |
| Youngstown-Warren-Boardman, OH-PA | East North Central | 7.40 | 17.92 | 20.20 | 20.52 | 25.52 | 27.06 | 30.01 | 3.57 | 4.93 | 22.61 | 4.99 |

\*Delta is the absolute increase in temperature from the 50th percentile to the 95th percentile
Abbreviations: 25th: 25th percentile of the temperature distribution; 95th: 95th percentile of the temperature distribution; 99th: 99th percentile of the temperature distribution; IQR: Interquartile range temperature; Max: Maximum temperature; Min: Minimum temperature; SD: Standard deviation temperature

**Table E7.**
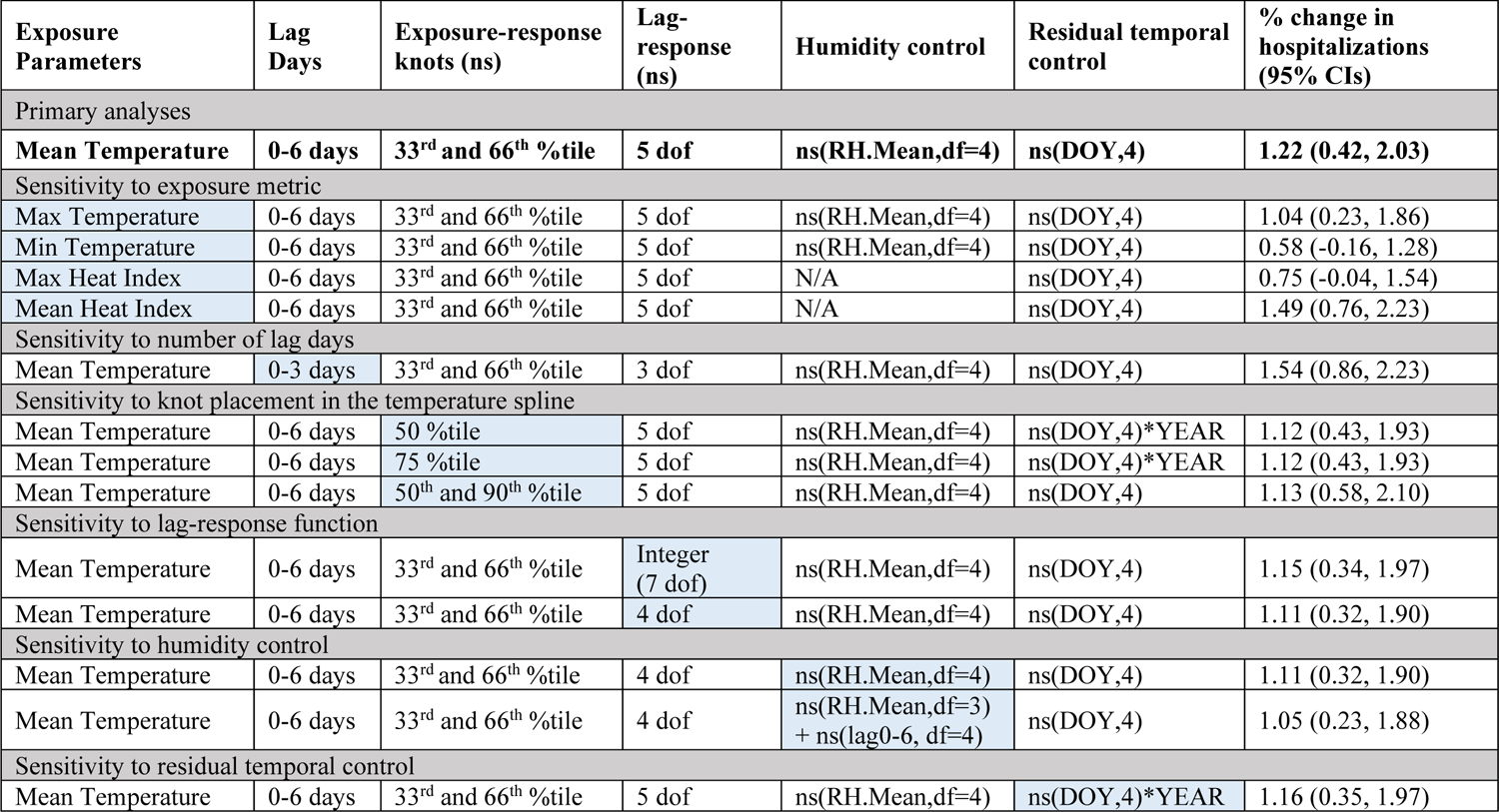
7-day cumulative, pooled relative risk (RR) from models performed under sensitivity analyses.

| Exposure Parameters | Lag Days | Exposure-response knots (ns) | Lag-response (ns) | Humidity control | Residual temporal control | % change in hospitalizations (95% CIs) |
| --- | --- | --- | --- | --- | --- | --- |
| Primary analyses |  |  |  |  |  |  |
| Mean Temperature | 0-6 days | 33<sup>rd</sup> and 66<sup>th</sup> %tile | 5 dof | ns(RH.Mean,df=4) | ns(DOY,4) | 1.22 (0.42, 2.03) |
| Sensitivity to exposure metric |  |  |  |  |  |  |
| Max Temperature | 0-6 days | 33<sup>rd</sup> and 66<sup>th</sup> %tile | 5 dof | ns(RH.Mean,df=4) | ns(DOY,4) | 1.04 (0.23, 1.86) |
| Min Temperature | 0-6 days | 33<sup>rd</sup> and 66<sup>th</sup> %tile | 5 dof | ns(RH.Mean,df=4) | ns(DOY,4) | 0.58 (-0.16, 1.28) |
| Max Heat Index | 0-6 days | 33<sup>rd</sup> and 66<sup>th</sup> %tile | 5 dof | N/A | ns(DOY,4) | 0.75 (-0.04, 1.54) |
| Mean Heat Index | 0-6 days | 33<sup>rd</sup> and 66<sup>th</sup> %tile | 5 dof | N/A | ns(DOY,4) | 1.49 (0.76, 2.23) |
| Sensitivity to number of lag days |  |  |  |  |  |  |
| Mean Temperature | 0-3 days | 33<sup>rd</sup> and 66<sup>th</sup> %tile | 3 dof | ns(RH.Mean,df=4) | ns(DOY,4) | 1.54 (0.86, 2.23) |
| Sensitivity to knot placement in the temperature spline |  |  |  |  |  |  |
| Mean Temperature | 0-6 days | 50 %tile | 5 dof | ns(RH.Mean,df=4) | ns(DOY,4)*YEAR | 1.12 (0.43, 1.93) |
| Mean Temperature | 0-6 days | 75 %tile | 5 dof | ns(RH.Mean,df=4) | ns(DOY,4)*YEAR | 1.12 (0.43, 1.93) |
| Mean Temperature | 0-6 days | 50<sup>th</sup> and 90<sup>th</sup> %tile | 5 dof | ns(RH.Mean,df=4) | ns(DOY,4) | 1.13 (0.58, 2.10) |
| Sensitivity to lag-response function |  |  |  |  |  |  |
| Mean Temperature | 0-6 days | 33<sup>rd</sup> and 66<sup>th</sup> %tile | Integer (7 dof) | ns(RH.Mean,df=4) | ns(DOY,4) | 1.15 (0.34, 1.97) |
| Mean Temperature | 0-6 days | 33<sup>rd</sup> and 66<sup>th</sup> %tile | 4 dof | ns(RH.Mean,df=4) | ns(DOY,4) | 1.11 (0.32, 1.90) |
| Sensitivity to humidity control |  |  |  |  |  |  |
| Mean Temperature | 0-6 days | 33<sup>rd</sup> and 66<sup>th</sup> %tile | 4 dof | ns(RH.Mean,df=4) | ns(DOY,4) | 1.11 (0.32, 1.90) |
| Mean Temperature | 0-6 days | 33<sup>rd</sup> and 66<sup>th</sup> %tile | 4 dof | ns(RH.Mean,df=3) + ns(lag0-6, df=4) | ns(DOY,4) | 1.05 (0.23, 1.88) |
| Sensitivity to residual temporal control |  |  |  |  |  |  |
| Mean Temperature | 0-6 days | 33<sup>rd</sup> and 66<sup>th</sup> %tile | 5 dof | ns(RH.Mean,df=4) | ns(DOY,4)*YEAR | 1.16 (0.35, 1.97) |

**Table E8:**
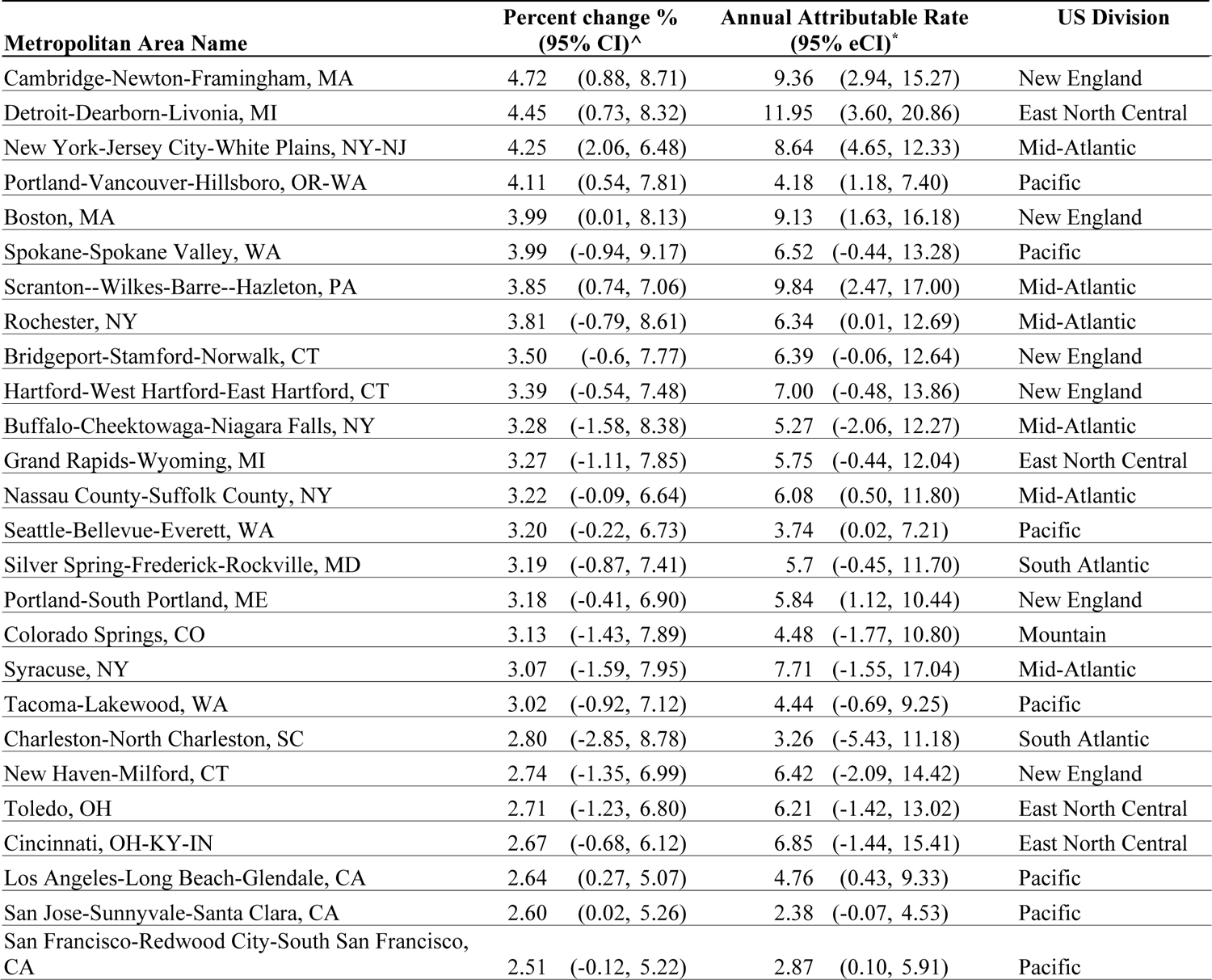
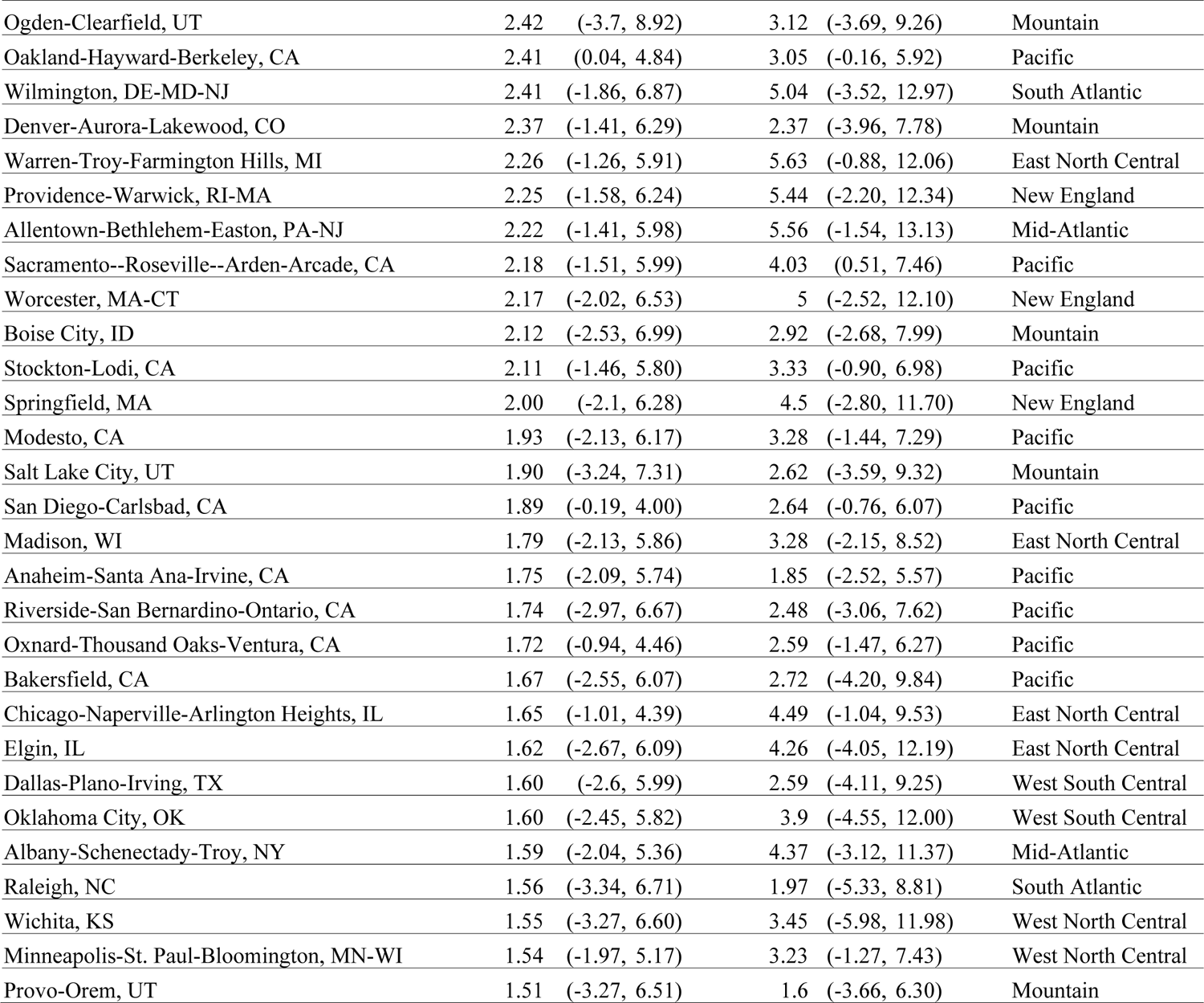
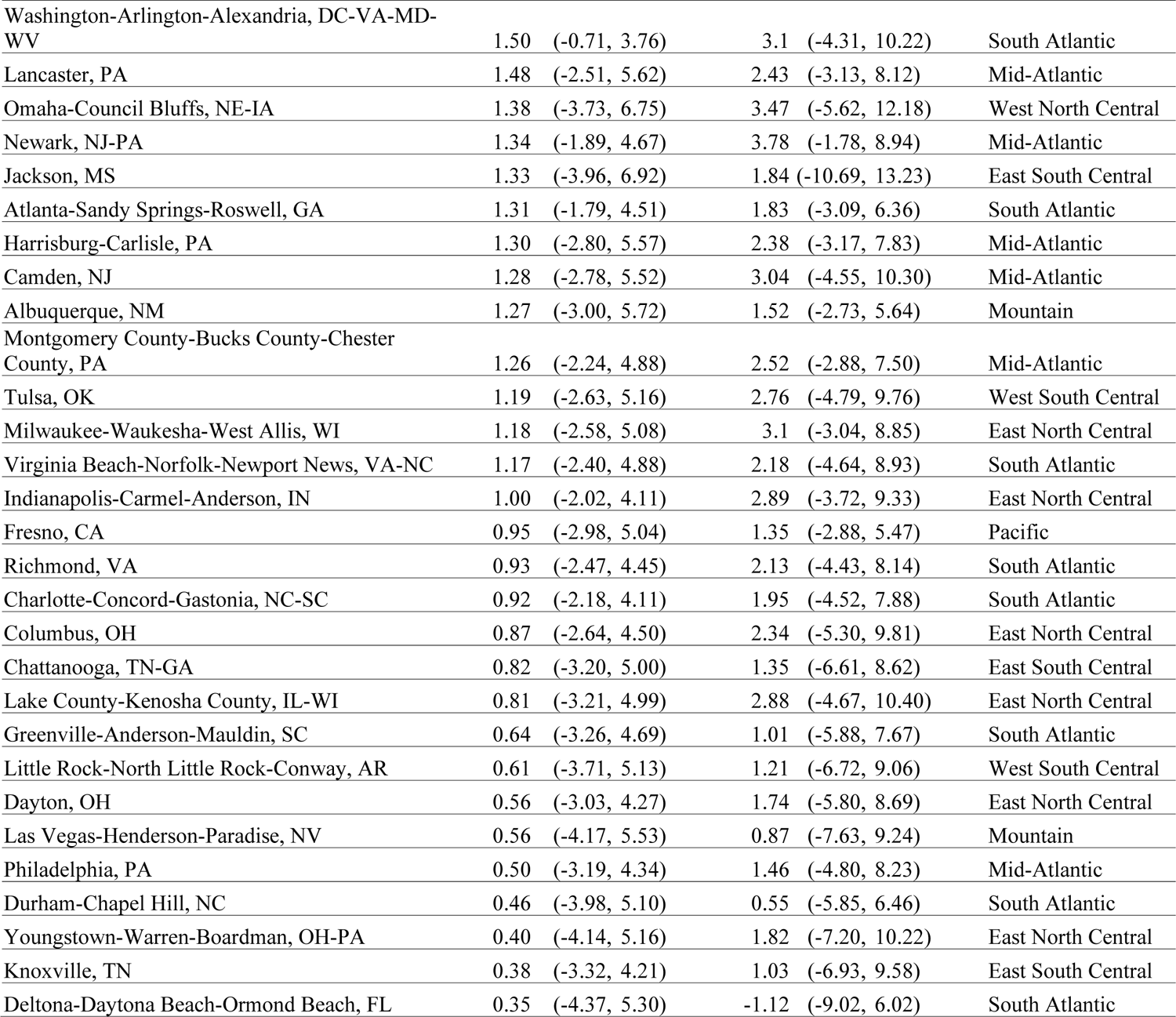
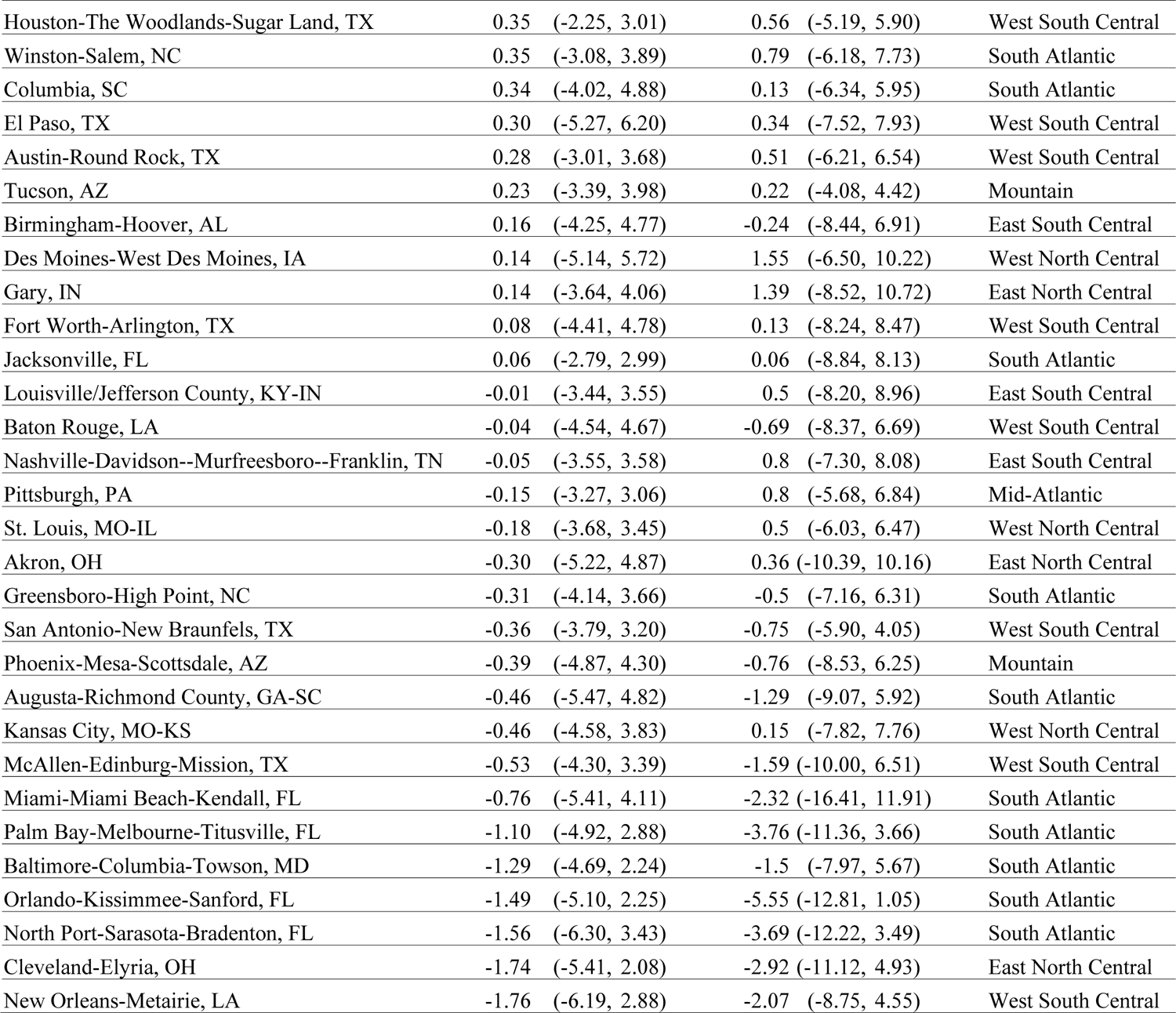
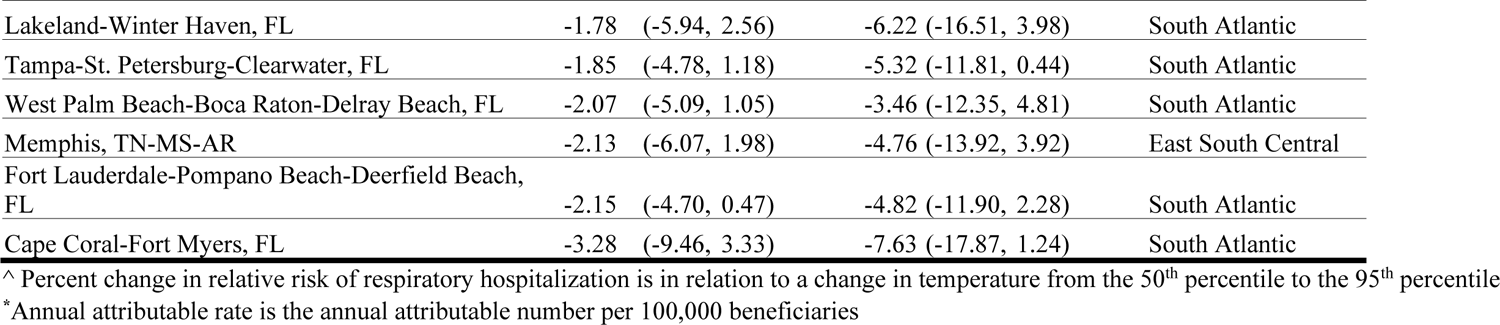
7-day cumulative percent change (95% CI) and annual attributable rate (95% CI) are reported for associations between high temperature and all-cause respiratory hospitalizations for each metropolitan area, 2000-2017.

